# Air cleaners and respiratory infections in schools: A modeling study using epidemiological, environmental, and molecular data

**DOI:** 10.1101/2023.12.29.23300635

**Authors:** Nicolas Banholzer, Philipp Jent, Pascal Bittel, Kathrin Zürcher, Lavinia Furrer, Simon Bertschinger, Ernest Weingartner, Alban Ramette, Matthias Egger, Tina Hascher, Lukas Fenner

## Abstract

**Background:** Using a multiple-measurement approach, we examined the real-world effectiveness of portable HEPA-air filtration devices (air cleaners) in a school setting.

**Methods:** We collected environmental (CO_2_, particle concentrations), epidemiological (absences related to respiratory infections), audio (coughing), and molecular data (bioaerosol and saliva samples) over seven weeks during winter 2022/2023 in two Swiss secondary school classes. Using a cross-over study design, we compared particle concentrations, coughing, and the risk of infection with vs without air cleaners.

**Results:** All 38 students (age 13*−*15 years) participated. With air cleaners, mean particle concentration decreased by 77% (95% credible interval 63%*−*86%). There were no differences in CO_2_ levels. Absences related to respiratory infections were 22 without vs 13 with air cleaners. Bayesian modeling suggested a reduced risk of infection, with a posterior probability of 91% and a relative risk of 0.73 (95% credible interval 0.44*−*1.18). Coughing also tended to be less frequent (posterior probability 93%). Molecular analysis detected mainly non-SARS-CoV-2 viruses in saliva (50/448 positive), but not in bioaerosols (2/105 positive) or HEPA-filters (4/160). The detection rate was similar with vs without air cleaners. Spatiotemporal analysis of positive saliva samples identified several likely transmissions.

**Conclusions:** Air cleaners improved air quality, showed a potential benefit in reducing respiratory infections, and were associated with less coughing. Airborne detection of non-SARS-CoV-2 viruses was rare, suggesting that these viruses may be more difficult to detect in the air. Future studies should examine the importance of close contact and long-range transmission, and the cost-effectiveness of using air cleaners.

## Introduction

Transmission of respiratory infections such as SARS-CoV-2 and influenza are difficult to mitigate and control. Person-to-person transmission occurs primarily through the release of respiratory particles containing the viruses. Recently, the focus has been on small respiratory particles called aerosols, which have been found to carry the majority of viruses during respiratory activities.^1^ Unlike larger respiratory droplets, which tend to settle quickly, aerosols can remain suspended in the air for several hours and travel long distances.^2^

Improved ventilation systems are critical for a healthy indoor environment and can reduce the risk of respiratory transmission,^3, 4^ especially in schools where students spend most of their time indoors during the week. Portable HEPA-air filtration devices (air cleaners) may be another cost-effective alternative to upgrading ventilation systems, but their impact on respiratory viral transmission is less clear. A population-level study reported a lower incidence of SARS-CoV-2 in US elementary schools using different ventilation strategies, including air filtration devices.^5^ While several studies showed that air cleaners reduce particle concentrations,^6–8^ an association with viral RNA load in airborne samples could not be found,^9, 10^ although recent studies showed that air cleaners effectively removed SARS-CoV-2 bioaerosols in hospitals and other indoor settings.^11–13^ Simulation studies have further demonstrated the efficacy of air cleaners in reducing the risk of indoor transmission of SARS-CoV-2,^14^ and other respiratory viruses.^15^ However, most simulation studies assume that the detection of RNA equals transmissible virus, despite recent data showing a relevant loss of viral infectivity in respiratory particles over time.^16^ To date, it remains unclear whether reducing particle concentrations and removing bioaerosols will reduce indoor transmission of respiratory infections.

We used a multiple-measurement approach to study transmission of respiratory viruses under non-pandemic conditions and the effect of air cleaners in a school setting with a cross-over study design in the winter of 2022/2023. We collected environmental data (CO_2_, particle concentrations), epidemiological data (absences likely related to respiratory infections), audio recordings (coughing), and molecular data (detection of viruses in bioaerosol and saliva samples) during a seven-week study period from January to March 2023 in two Swiss secondary school classes. We determined changes in particle concentrations, absences related to respiratory infections, coughing, and the rate of positive saliva samples.

## Methods

This study is reported as per the Strengthening the Reporting of Observational Studies in Epidemiology guideline (see STROBE checklist).

### Study setting and design

We collected data in two classrooms of a secondary school (age of students 14-17 years) in the canton of Solothurn, Switzerland, for seven weeks from January 16 to March 11, 2023. Figure 1 shows the schematic study setup.

**Fig 1.**
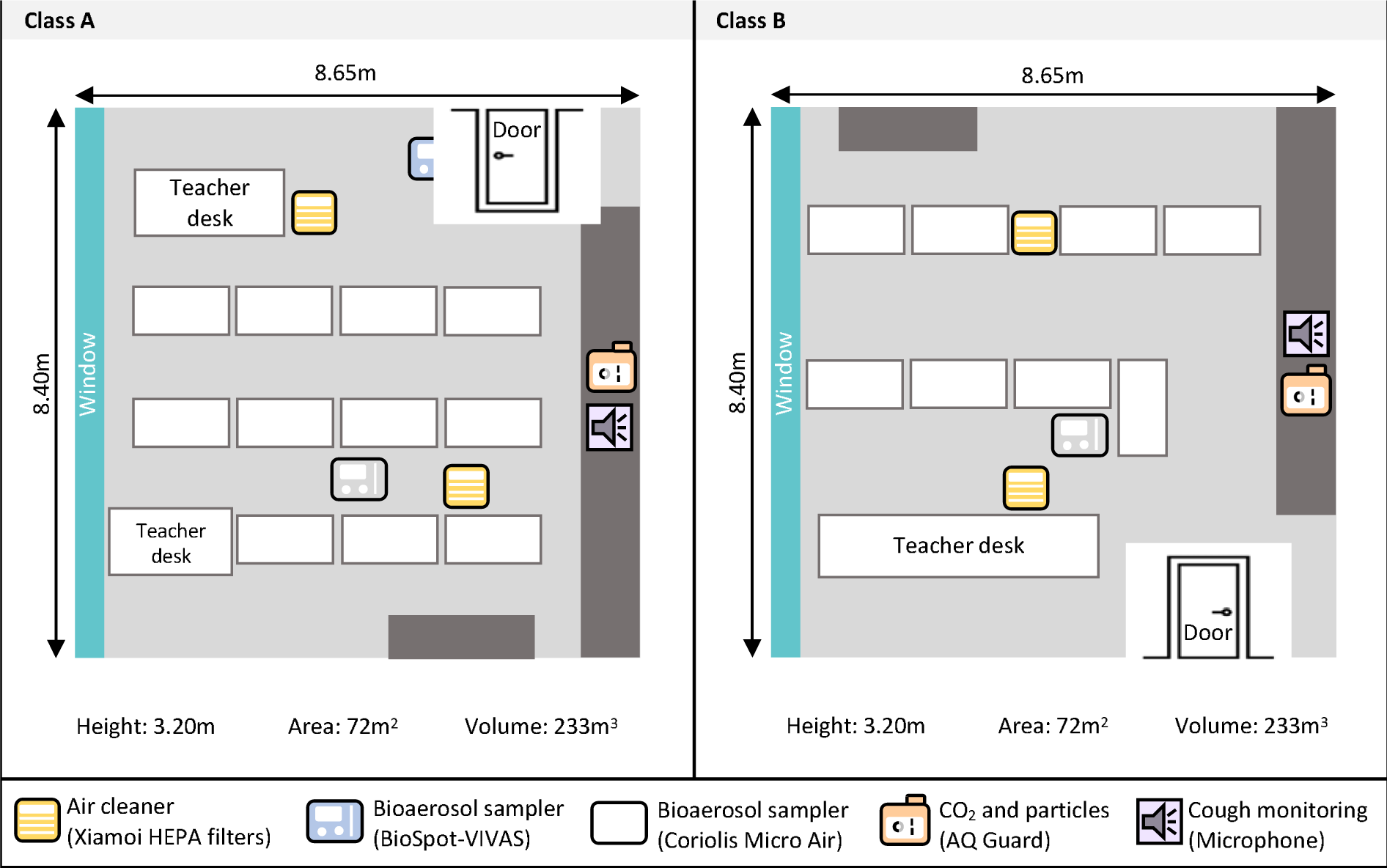
Study setting. Schematic study setup of the classrooms. One air cleaner was placed in the front and one in the back of the classrooms. All devices were placed at the head level of the students when they were seated. Both classrooms lacked an active HVAC (Heating, Ventilation, Air conditioning) system, but they were ventilated naturally by opening windows at the discretion of the teachers.

### Study intervention

We used a cross-over design to study the effectiveness of air cleaners (Table 1). Air cleaners refer to commercially available portable HEPA-filtration devices (Xiaomi Mi Air Pro 70m2, Shenzhen, China). According to the manufacturer, these air cleaners run at clean air delivery rates of 2 *×* 600 m^3^/h. When testing the devices in an empty classroom with sub-micrometer sized particles, we measured a lower effective clean air delivery rate of 2 *×* 420 m^3^/h (Supplementary Text A).

**Table 1.** Cross-over study design. Description of when portable air cleaners (PAC) were installed in the rooms of classes A and B during a seven-week study period from January 16 to March 11, 2023, excluding a week of vacation from February 6 to 11.

|  | **Week 1**<br>Jan 16-21 | **Week 2**<br>Jan 23-28 | **Week 3**<br>Jan 30-Feb 3 | **Vacation**<br>Feb 6-11 | **Week 4**<br>Feb 13-18 | **Week 5**<br>Feb 20-25 | **Week 6**<br>Feb 27-Mar 3 | **Week 7**<br>Mar 6-11 |
| --- | --- | --- | --- | --- | --- | --- | --- | --- |
| **A** | PAC | PAC * | None |  | None | None | PAC | PAC * |
| **B** | None | None | PAC * |  | PAC | PAC * | None | None |

\* Swabs from the HEPA filters taken after each intervention phase and before the vacation.

### Data collection

An overview of the collected data is provided in Supplementary Text B and Supplementary Table S1.

#### Environmental data

An air quality device (AQ Guard, Palas GmbH, Karlsruhe, Germany) continuously measured indoor CO_2_ levels, aerosol number (particle diameter between 175 nm to 20 *µ*m) and particle mass concentrations (PM; PM_1_, PM_2.5_, PM_4_, PM_10_) by minute.^8^

#### Epidemiological data

At study start, we collected aggregated data on age, sex, COVID-19 vaccination and recovery status in the participating classes. Daily, we collected data on each absent student. For absences due to illness, we recorded symptoms and the date of symptom onset. We defined a case of respiratory infection as an absence in which the student reported an illness with at least one respiratory symptom (Supplementary Text C). All absences are listed in Supplementary Table S2.

### Audio recordings and cough detection

We installed portable audio recorders (ZOOM H6; New York, USA) to record sounds continuously. We determined the number of coughs per minute using an AI algorithm.^17^ A recent study showed a significant correlation between coughing and airborne viral detection.^10^

#### Molecular data analyses

Both classes participated in repetitive bi-weekly (Tuesdays and Thursdays) saliva testing. Samples were transported to the laboratory and stored at *−*80°C until further processing.^18^ All positive samples are listed in Supplementary Table S3 and Text D. Furthermore, we collected airborne respiratory viruses in both classrooms with a cyclonic bioaerosol sampling device (Coriolis Micro Air, Bertin Instruments Montigny-le-Bretonneux, France), running at 200 l/min and collecting into 15 mL Phosphate-Buffered Saline. The Coriolis Micro Air ran shortly before and during break times (approximately 60 min/day) to minimize noise. In one class, we also sampled with the BioSpot-VIVAS condensation particle growth collection device (Aerosol Devices Inc., Ft. Collins, CO, USA),^19^ which operated throughout lessons. The removable parts were regularly autoclaved. At the end of the day, samples were transported to the Institute of Infectious Diseases and stored at *−*80°C. Finally, we collected swabs from the air cleaners’ HEPA filters after each intervention phase (see Table 1). The HEPA filters were removed and divided into 20 fields. One sterile Phosphate-Buffered Saline-moistened swab per field was then taken for a total of 20 swabs per filter.

Prior to the real-time (RT)-PCR analysis, daily bioaerosol samples were combined for each sampling device and filtered using Amicon Ultra-15 Centrifugal Filters with Ultracel 10,000 Dalton molecular weight cutoffs filters (UFC9010; MilliporeSigma, Burlington, USA) to a volume of 1 mL. Saliva samples were analyzed directly without prior filtration. The Allplex RV Master Assay (Seegene, Seoul, South Korea) detects a panel of 19 major respiratory viruses and viral subtypes, including SARS-CoV-2, influenza, respiratory syncytial, metapneumovirus, adenovirus, rhinovirus, and parainfluenza.

We also performed molecular genotyping for positive saliva, bioaerosol, and air filter samples of adenovirus and influenza.^20^

### Statistical analyses and modeling

All statistical analyses were described in a statistical analysis plan^21^. Bioaerosol samples and viral load concentrations could not be analyzed because there were too few positive samples. Further minor deviations from the statistical analysis plan are documented in Supplementary Texts E*−*H, including detailed descriptions of the models.

#### Particle concentrations

We compared daily mean particle concentrations between study conditions (Supplementary Figure S1). We estimated the reduction in particle concentrations with air cleaners using Bayesian log-linear regression models, adjusting for observed confounders (Supplementary Text E).

#### Risk of infection

We estimated the relative risk of infection with air cleaners using a Bayesian latent variable regression model (Supplementary Text F). The number of new respiratory cases *C* (observed absences related to respiratory infections by date of symptom onset) on day *t* in class *j* are modeled with a Negative Binomial distribution. The expected number of new cases is the weighted sum of the number of new infections *I_js_* (latent variable) in the previous days *s < t*, with the weights corresponding to the probability distribution of the incubation period (Supplementary Figure S2). The number of new infections is related to the presence of air cleaners as follows:

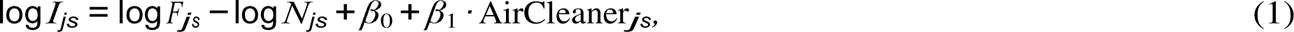

where *F_js_* is the number of infections in the previous week (a proxy for the number of infectious students), *N_js_* is the cumulative number of infections (a proxy for the number of susceptible students), β_0_ is the infection rate without air cleaners, and β_1_ is the effect of air cleaners. Furthermore, the effect of air cleaners is adjusted for class-specific effects, the number of students in class, the daily air change rate, and the weekly positivity rate for COVID-19 and the consultations for influenza-like illnesses in the canton.

#### Coughing

We estimated the reduction in the daily number of coughs with air cleaners using a Bayesian Negative Binomial regression model, using time in class as the model offset and adjusting for observed confounders (Supplementary Text G). In addition, we estimated the association between the number of coughs and the virus-specific number of positive saliva samples, using only the days when saliva samples were collected (Tuesdays and Thursdays, for a total of 27 days).

#### Saliva samples

We analyzed the number of positive saliva samples with a Bayesian Multinomial logistic regression model (Supplementary Text H) and linked the expected number of positive samples to the presence of air cleaners, adjusting for the cumulative number of positive tests.

#### Software

All analyses were performed in R software (version 4.2.0) and model parameters estimated in Stan (version 2.21.0).^22, 23^ For each outcome, we report the posterior probability of a reduction with air cleaners. The estimated reduction is reported with the posterior mean and 95%credible interval (CrI).

### Ethics statement

The Ethics Committee of the canton of Bern, Switzerland, approved the study (reference no. 2021-02377). For the saliva samples, we included all students who were willing to participate and obtained written informed consent from their caregivers.

## Results

The study population consisted of 38 students (age 13*−*15 years, 19/19 female/male; Table 2). Seven students had been vaccinated or recovered from a SARS-CoV-2 infection within the last four months. During the seven-week study period (total of 1,330 student-days), students were absent from school for 220 days (18% of the total) of which 129 days (59% of absences) were due to illness.

**Table 2.** Overview of the study population and person-days of absences.

|  | Class A | Class B | Total |
| --- | --- | --- | --- |
| **Students** | **20 (53%)** | **18 (47%)** | **38 (100%)** |
| *Gender* |  |  |  |
| - Female | 11 (55%) | 8 (44%) | 19 ( 50%) |
| - Male | 9 (45%) | 10 (56%) | 19 ( 50%) |
| *Immunity status* |  |  |  |
| - Recently vaccinated (or recovered) | 7 (39%) | 0 (0%) | 7 ( 18%) |
| - Not recently vaccinated (or recovered) | 11 (61%) | 20 (100%) | 31 ( 82%) |
| **Absent person-days** | **110 (50%)** | **110 (50%)** | **220 (100%)** |
| - Sickness | 52 (47%) | 77 (70%) | 129 ( 59%) |
| - Other | 58 (53%) | 33 (30%) | 91 ( 41%) |

### Air quality

The mean aerosol number concentration was 95 1/cm^3^ (standard deviation [SD] 81 1/cm^3^) without vs 27 1/cm^3^ (SD 34 1/cm^3^) with air cleaners (Figure 2a). The Bayesian regression model suggested a clear reduction in the aerosol concentration with air cleaners, with a posterior probability of 100%. The model-estimated decrease was 76% (95%-CrI 63% to 86%), which was greater for larger (PM_10_) than for smaller (PM_1*−*4_) particles (Supplementary Figure S3, Supplementary Table S4). Daily mean CO_2_ levels were comparable between study conditions (1,636 ppm (SD 341 ppm) without vs 1,769 ppm (SD 391 ppm) with air cleaners). There was little change in other environmental variables (Supplementary Figure S4).

**Fig 2.**
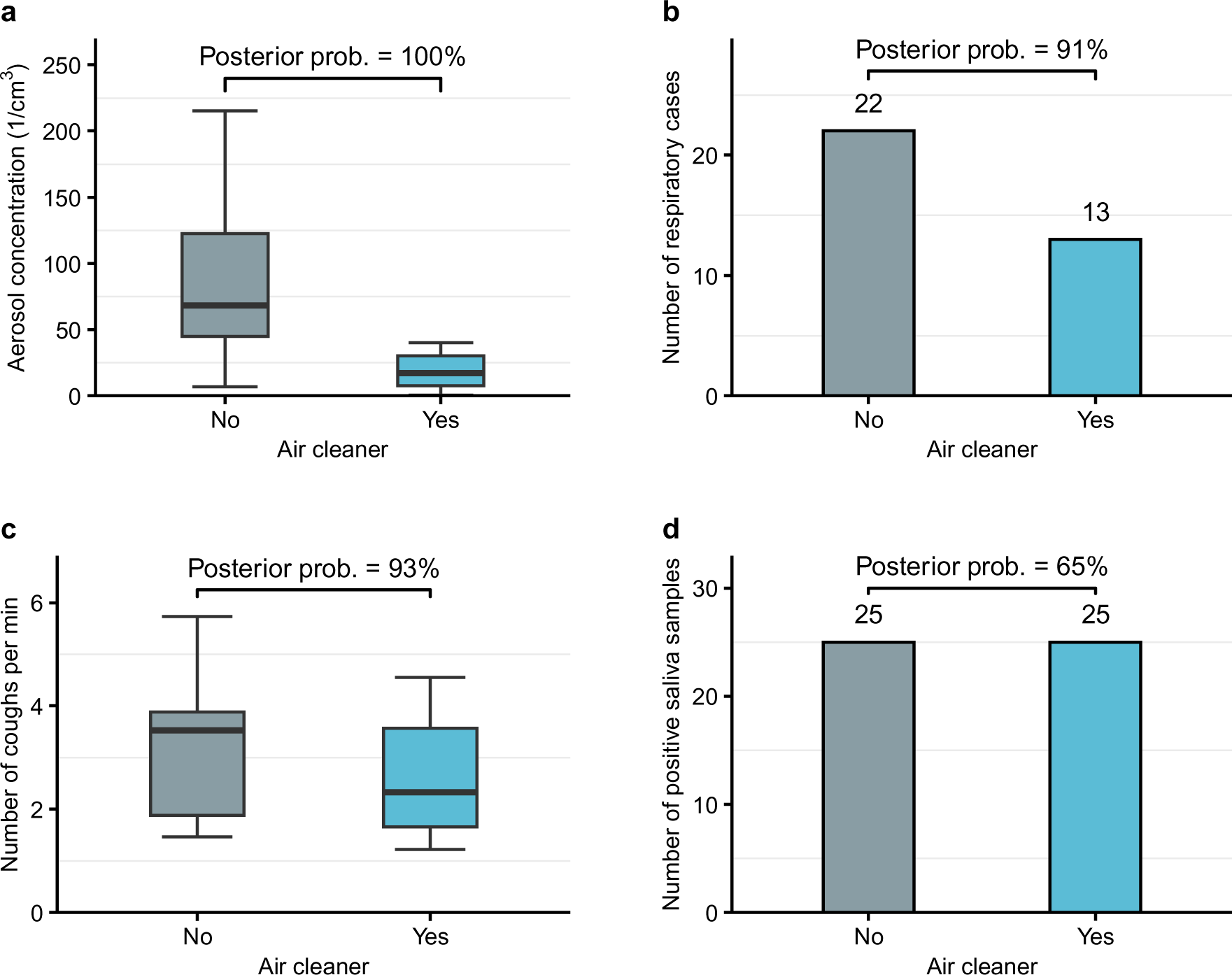
Comparison of outcomes with vs without air cleaners. At the top of each plot, the posterior probability for a reduction with air cleaners is shown, based on the Bayesian model. **(a)** Daily average aerosol number concentrations as boxplots. **(b)** Number of respiratory cases. **(c)** Daily average number of detected coughs per minute as boxplots. **(d)** Number of positive saliva samples.

### Risk of infection

Absences related to respiratory infections included 22 cases without vs 13 cases with air cleaners (Figure 2b). The Bayesian latent variable hierarchical regression model suggested that air cleaners reduced the risk of infection, with a posterior probability of 91%. The adjusted relative risk of infection with air cleaners was 0.73 (95%-CrI 0.44 to 1.18). The estimated number of respiratory infections in school would have been 19 (95%-CrI 9 to 37) if air cleaners had been installed throughout the study period, compared to 36 (95%-CrI 12 to 92) infections if air cleaners had not been installed. Detailed estimation results are provided in Supplementary Information (Supplementary Text J, Supplementary Figure S5, Supplementary Table S5).

### Coughing

On average, we detected 3.1 coughs/min (SD 1.2 coughs/min) without vs 2.6 coughs/min (SD 1.1 coughs/min) with air cleaners (Figure 2c). The Bayesian model suggested that coughing was less frequent with air cleaners, with a posterior probability of 93%. The adjusted relative risk of coughing with air cleaners was 0.93 (95%-CrI 0.85 to 1.02). Coughing was associated with virus-specific transmission (Supplementary Figure S6).

### Saliva samples

We analyzed a total of 448 saliva samples. We detected 15 influenza B, 15 rhinovirus, 14 adenovirus, 3 SARS-CoV-2, 2 metapneumovirus, and 1 parainfluenza virus, respectively (Figure 3a). There were 25 positive saliva samples in both study conditions (Figure 2d) and, based on the Bayesian model, the posterior probability that air cleaners reduced the positivity rate was 65%, with an adjusted relative risk of 0.93 (95%-CrI 0.49 to 1.61). The estimated relative risk was not sensitive to infection to testing delays (Supplementary Figure S7).

**Fig 3.**
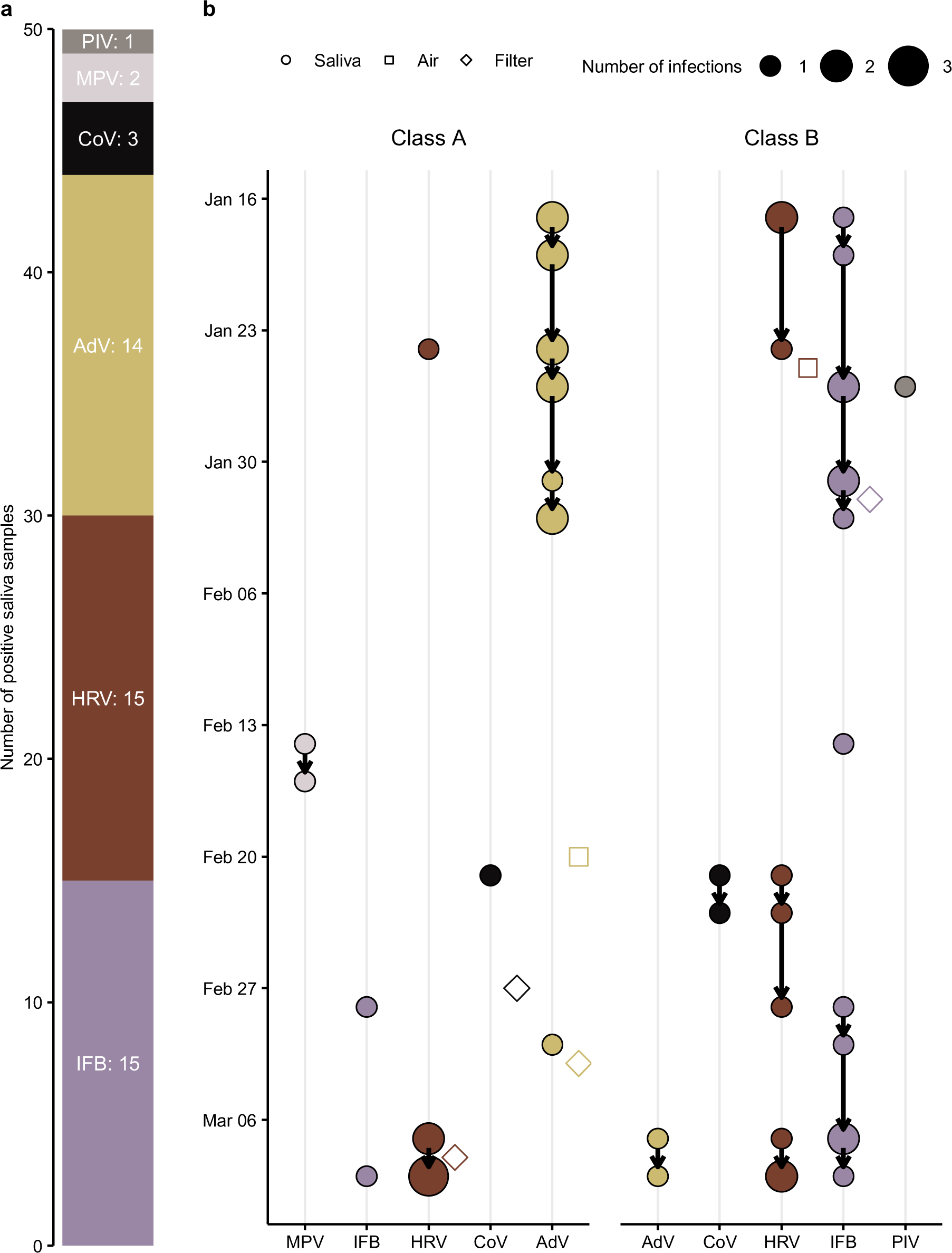
Molecular detection of respiratory viruses and transmission network based on spatiotemporal analysis of students’ saliva samples. **(a)** Number of positive saliva samples by virus. **(b)** Daily number of positive saliva samples (colored circles) and possible transmission chains within classes (directed arrows). Positive samples are linked if they belong to the same virus and are less than 1 week apart. Positive samples from the air and filters as blank squares aligned. IFB: influenza B, HRV: human rhinovirus, AdV: adenovirus, CoV: SARS-CoV-2, MPV: human metapneumovirus, PIV: parainfluenza virus.

### Transmission patterns

The distribution of positive saliva samples varied between classes. For example, all but one sample was positive for adenvovirus in class A during the first three study weeks, while the vast majority of positive influenza B samples were found in class B. To illustrate possible transmission chains within classes, we linked positive saliva samples of the same virus that were less than one week apart. Based on this spatiotemporal analysis, we identified 10 possible transmission chains (Figure 3b). The longest potential transmission chains occurred in January, referring to a cluster of adenovirus infections in class A and influenza B infections in class B. Molecular genotyping to verify the proposed transmission network was unsuccessful because we could not amplify and sequence any of the gene targets. We also analyzed 105 bioaerosol samples and detected in two of them viral RNA (1 rhinovirus in class A and 1 adenovirus in class B). Similarly, we detected 1 influenza B, 1 rhinovirus, 1 adenovirus, and 1 SARS-CoV-2 in the 20 swabs taken from each filter of an air cleaner after each intervention phase (160 swabs in total).

## Discussion

We used a multiple-measurement approach within a cross-over study design to estimate the risk of respiratory virus infection in a Swiss school and to assess the effectiveness of air cleaners. We found a wide range of respiratory viruses in saliva samples, mainly adenovirus, influenza B, and rhinovirus, but very few viral RNA was detected in bioaerosol samples and on the filters of the air cleaners. Particle mass concentrations decreased significantly with air cleaners, and Bayesian modeling based on epidemiological data indicated a reduction in the relative risk of infection with air cleaners. Coughing was reduced, compatible with air cleaners preventing some symptomatic infections.

We detected a range of respiratory virus in students’ saliva, mainly adenovirus, influenza B and rhinovirus, with only three positive SARS-CoV-2 saliva samples. In a previous, similar study in the same setting, we estimated the effectiveness of mask wearing and air cleaners during the SARS-CoV-2 omicron wave in the winter of 2021/2022 and detected almost exclusively SARS-CoV-2 in the students’ saliva.^8^ A similar shift in the pattern of respiratory viruses has been observed in other studies.^24, 25^ We found a reduction in the risk of SARS-CoV-2 infection for mask wearing, but not for air cleaners, possibly because the air cleaners were introduced only at the end of the study, when most students were already infected with SARS-CoV-2.

It is well documented that air cleaners improve indoor air quality.^6–8^ There are several reasons why the effect of air cleaners is probably smaller than universal mask wearing, which has been shown to be a very effective infection control measure.^5, 8, 26, 27^ Unlike masks, air cleaners cannot prevent transmission outside the classroom or transmission due to close range, high particle density. Prolonged and close contact may be necessary for transmission of some respiratory viruses,^26, 28^ or make transmission more likely despite prior vaccination or infection.^29^ Our results further suggests that air cleaners are more effective at removing larger particles (*>* 5*µ*m), which also explains the difference between our measured and the manufacturer’s reported clean air delivery rate (see Supplementary Text A). However, many respiratory viruses are carried in smaller particles, which are more relevant for transmission (≤ 5*µ*m).^1^ Finally, classroom activity, airflow and other unobserved, confounding factors make it challenging to evaluate the effects of air cleaners on transmission in real-world settings.

The beneficial effects of air cleaners on indoor air quality and transmission come at a reasonable cost. The portable air cleaners used in our study cost approximately USD 250 per unit. Their operating cost-effectiveness in providing clean air could be even higher than that of a ventilation system when compared in parallel using the same air delivery ratings.^30^ Therefore, air cleaners could be a cost-effective public health measure, particularly during pandemics or epidemics when there is greater exposure to respiratory infections, and greater concern of becoming infected.^31^ However, their acceptance may be hindered by noise, space limitations, technical issues, and maintenance requirements.^32^ Therefore, investments in professional building ventilation systems are still preferred in the long run.^33^

We detected only few respiratory viruses in bioaerosol samples (1 sample of adenovirus and 1 sample of rhinovirus) and on the filters of the air cleaners (4 positive samples). The low rate of positive bioaerosol samples may indicate that it is unlikely that airborne transmission occurred in classrooms. It may be possible that students had relatively little exposure to respiratory viruses at school and acquired their infections elsewhere. However, the distribution of positive saliva samples markedly differed between classes. Adenovirus spread in class A during the first three study weeks, with only two infections of influenza B over the study period. In contrast, influenza B spread throughout the study in class B, and adenovirus infections were detected only in the last week of the study. Furthermore, adenovirus infections tend to be mild,^34^ and less frequently associated with cough than influenza,^35^, consistent with the comparatively lower frequency of coughing in class B. Taken together, the class-specific, spatiotemporal patterns indicate that transmission of respiratory infections may have occurred within the classrooms.

Our study has limitations. Aerosol measurements and molecular detection of viruses in bioaerosol samples document exposure, but not transmission and the direction of transmission (person to air, air to person) cannot be determined. Further, the reasons for school absences were self-reported by students and some absences may have been incorrectly attributed to respiratory infections. In addition, we could only approximate the incubation period for each epidemiological case. Finally, although the study results are likely to apply to many settings in Switzerland and other European countries, they will not be applicable to settings in the global South.

In conclusion, a wide range of respiratory viruses, but rarely SARS-CoV-2, was detected in students under non-pandemic conditions when public health measures were lifted. Airborne detection was rare, suggesting that respiratory viruses other than SARS-CoV-2 may be more difficult to detect and that prolonged close contact may be required for transmission. The risk reduction of respiratory infections conferred by air cleaners may be modest at the individual level, but the benefit at the population level in terms of illness and absences prevented is likely to be important. Future studies should examine the cost-effectiveness of using air cleaners in congregate settings.

## Acknowledgements

We would like to thank the schools, teachers, and students participating in the study. We are grateful to the Educational Department of the canton of Solothurn for their support throughout the study. We would also like to thank Ronald Dijkman for the influenza molecular genotyping and the biosecurity team (Julia Feldmann, Monika Gsell, Kathrina Summermatter) from the Institute for Infectious Diseases at the University of Bern for their assistance with the bioaerosol devices. Finally, we are indebted to the student assistants (Khadija Ahmed, Marie-Joséphine Brancato, Michelle Bürki, Santiago Martinez, Moric Toszeghi, Sylvain Wasmer) who helped with the data collection in the schools.

## Data availability

Preprocessed data and code are available from https://osf.io/38j9g.

## Funding

This work was supported by the Multidisciplinary Center for Infectious Diseases, University of Bern, Switzerland. NB, LF, and ME are supported by the National Institute of Allergy and Infectious Diseases (NIAID) through cooperative agreement 5U01-AI069924-05. ME is supported by special project funding from the Swiss National Science Foundation (grant 32FP30-189498). The funder of the study had no role in study design, data collection, data analysis, data interpretation, or writing of the report.

## Author contributions

Conception and design: NB, KZ, LF, PB, PJ, TS. Epidemiological and environmental data collection: EW, NB, PJ, KZ, TS, LF. Laboratory data collection: PB, LFu. Additional data collection: TH. Cough detection: SB. Molecular genotyping: AR, PB, LFu. Statistical analysis: NB. Paper draft: NB, LF, ME. All authors reviewed and approved the final version of the manuscript.

## Notes

### Competing Interest Statement

The authors have declared no competing interest.

### Author Declarations

The Ethics Committee of the canton of Bern, Switzerland, approved the study (reference no. 2021- 02377). For the saliva samples, we included all students who were willing to participate and obtained written informed consent from their caregivers.

