## Supplementary Information for "Air cleaners and respiratory infections in schools: A modeling study using epidemiological, environmental, and molecular data"

### Supplmenetary Material

### List of Texts

**A Experimental determination of the CADR** **4**

**B Data collection** **6**

**C Epidemiological line list data** **7**

**D Molecular line list data** **9**

**E Modeling changes in particle concentrations** **22**

**F Modeling relative risk of infection** **23**

**G Modeling changes in cough frequency** **28**

**H Modeling positivity rate of human saliva samples** **29**

**I Detailed results for changes in particle concentrations** **30**

**J Detailed results for risk of infection model** **32**

**K Detailed results for cough frequency** **35**

**L Detailed results for positivity rate in saliva samples** **36**

**References** **37**

### A Experimental determination of the CADR

The clean air delivery rate (CADR) is typically used to quantify the cleaning efficiency of air cleaners.[1] It is expressed in $m^3/s$ and describes the volumetric flow rate of particle-free, clean air that an air cleaner delivers into the room. The CADR is equal to the product of the filtration efficiency and the volumetric flow rate passing through the device.

We followed the approach by Küpper et al. to determine the CADR.[2] The approach is based on the release of fine aerosols (particle diameter $D<1\mu$m) in a closed, unventilated room by measuring the decay rate $k$ of the particle concentration $N(t)$ caused by the air cleaner, i. e.

$$N(t) = N_0 \cdot e^{-k \cdot t} \tag{1}$$

Since the surfaces in the room (e. g. walls and furniture) are also sinks for aerosol particles, measurements were carried out with ($k_{\text{cleaner}}$) and without air cleaners($k_{\text{background}}$). Based on that, we can compute the CADR as

$$\text{CADR} = V \cdot k_{\text{cleaner}} - V \cdot k_{\text{background}}, \tag{2}$$

where $V = 233m^3$ is the volume of the classroom.

We ensured that the particle concentrations in the experiments were not too high, thereby preventing bias due to coagulation (i. e. the formation of a few large particles from many small particles). At the beginning of each experiment, fine aerosol particles were released for a few minutes by ultrasonic nebulization of an aqueous NaCl solution of 5 g/l. The generated dehydrated salt particles have a broad size distribution in the range of $0.02-1.0\mu$m. Decay rates were measured using 11 fine dust sensors (SPS30, Sensirion AG), which were placed at head height of the students. These sensors measured the number concentration of individual particles by counting the scattered light pulses they produce when passing through a laser beam. Because of their scattering properties, these sensors measure the number of particles larger than $\sim 0.3\,\mu$m.

Figure S1 shows the particle number concentration with and without without the air cleaner. The exponential decrease is clearly visible as a linear curve in the logarithmic plot. Based on these experiments, we were able to determine the net CADR values of the air cleaner as $420\,m^3/h$

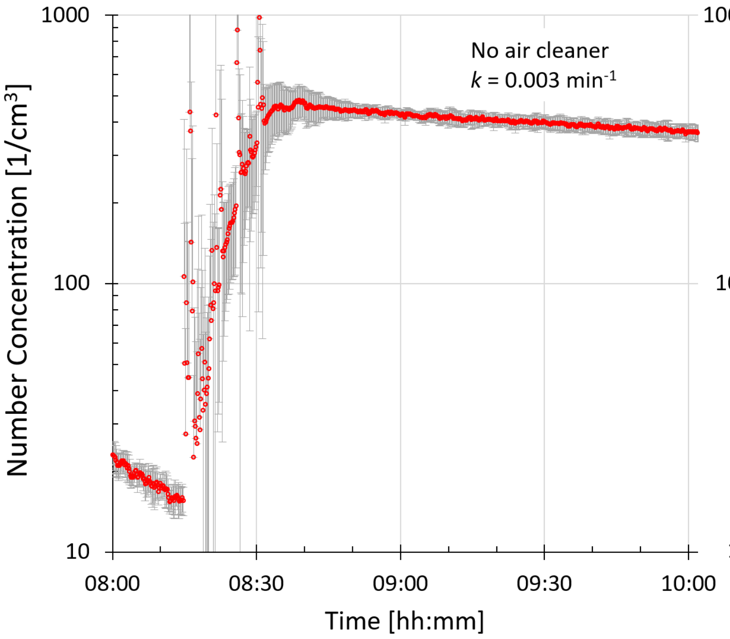

**Figure S1. Measured concentration curves with and without air cleaner.** The red dot shows the average concentration measured by the 11 distributed sensors of particles with $0.3\mu m < D < 1\mu m$. The standard deviation is shown in gray as an error bar. The increase in concentration at the beginning of these experiments is due to the release of the NaCl particles during approximately 15-20 min. The inhomogeneity of the concentration during the nebulization period is seen in larger error bars. Afterwards, the aerosol source is turned off, the concentration homogenizes quickly, and the concentration drop follows an exponential decay, which is used for the CADR calculation.

with an uncertainty of ±10% due to systematic differences in the setup and measurement errors of the instruments. Our estimated CADR is lower than the CADR reported by the manufacturer of $600 m^3/h$, most likely because the reported CADR was determined with comparably large particles (pollen in the super micrometer size range). Thus, we explain the lower filtration efficiency determined here with the fact that CADR is strongly dependent on the particle size, i. e. large particles are easier to filter than fine aerosols.

### B Data collection

Table S1 in SI Appendixprovides an overview of the types of data that were collected in each classroom.

**Table S1. Overview of collected data**. Type of data collected, method/device, and frequency in the rooms of classes A and B.

| Type of data | Method / device | Class A | Class B | Frequency |
|---|---|---|---|---|
| **Molecular data** | | | | |
| Bioaerosol sampling | Bioaerosol sampling devices: BioSpot-VIVAS (Aerosol Devices Inc., Ft. Collins, Colorado, United States) and Coriolis Micro Air (Bertin Instruments Montigny-le/Bretonneux, France) | x | (x) only Coriolis | daily |
| Saliva samples | Saliva samples | x | x | twice per week |
| Swabs from air cleaners | Swab samples from Xiaomi HEPA filters (Xiaomi Mi Air Pro 70m2, Shenzhen, China) | x | x | after each intervention phase (both classes) and before the vacation (only class B) |
| **Environmental data** | | | | |
| $CO_2$, aerosol/particle concentrations, humidity, temperature | Air quality device: AQ Guard (Palas GmbH, Karlsruhe, Germany) | x | x | daily by minute |
| Coughs | Sounds collected via microphones and coughs detected with a Machine Learning model[3] | x | x | daily by seconds |
| **Epidemiological data** | | | | |
| Absences | Survey | x | x | daily |
| Student characteristics | Survey | x | x | at study start |

### C Epidemiological line list data

Reports about absences were entered electronically into a REDcap database.[4,5] Table S2 shows the line list data for respiratory cases based on epidemiological data. For each class and case, it shows the date when the student was absent, s/he reported symptoms, and returned to school. We also collected data on laboratory tests but none of the students reported a test result during the study. A case of respiratory infection was defined as an absence where the student reported a sickness with at least one of the following symptoms: fever, coughing, tiredness, loss of test or smell, sore throat, headache, aches and pains, diarrhea, difficulty breathing or shortness of breath, stomach. Students were asked to report the first date they experienced symptoms, so that a student absent on Monday could report Saturday or Sunday as the day when symptoms began to be felt. We will refer respiratory cases by their dates of symptom onset, which usually corresponded to the absence date, unless the student attended the school while experiencing symptoms. Informed consent for one student could not be obtained and the single absence of this student was considered unrelated to a respiratory infection.

**Table S2.** Line list of respiratory cases over the study period.

| Class | Date of absence | Date of symptom onset | Date of return |
|---|---|---|---|
| A | 2023-01-16 | 2023-01-16 | 2023-01-19 |
| A | 2023-01-23 | 2023-01-23 | 2023-01-24 |
| A | 2023-02-05 | 2023-02-05 | 2023-02-13 |
| A | 2023-02-09 | 2023-02-09 | 2023-02-13 |
| A | 2023-02-17 | 2023-02-17 | 2023-02-20 |
| A | 2023-02-20 | 2023-02-20 | 2023-02-21 |
| A | 2023-02-20 | 2023-02-20 | 2023-02-23 |
| A | 2023-02-21 | 2023-02-21 | 2023-02-22 |
| A | 2023-02-27 | 2023-02-27 | 2023-02-28 |
| A | 2023-02-27 | 2023-02-27 | 2023-03-01 |
| A | 2023-02-25 | 2023-02-27 | 2023-03-06 |
| A | 2023-02-27 | 2023-02-28 | 2023-03-03 |
| A | 2023-03-06 | 2023-03-06 | 2023-03-07 |
| A | 2023-03-06 | 2023-03-06 | 2023-03-09 |
| B | 2023-01-16 | 2023-01-16 | 2023-01-17 |

| | | | |
|---|---|---|---|
| B | 2023-01-19 | 2023-01-19 | 2023-01-20 |
| B | 2023-01-16 | 2023-01-20 | 2023-01-23 |
| B | 2023-01-21 | 2023-01-23 | 2023-01-24 |
| B | 2023-01-23 | 2023-01-23 | 2023-01-24 |
| B | 2023-01-24 | 2023-01-24 | 2023-01-25 |
| B | 2023-02-01 | 2023-02-02 | 2023-02-13 |
| B | 2023-02-03 | 2023-02-03 | 2023-02-13 |
| B | 2023-02-17 | 2023-02-17 | 2023-02-21 |
| B | 2023-02-20 | 2023-02-20 | 2023-02-21 |
| B | 2023-02-20 | 2023-02-20 | 2023-02-28 |
| B | 2023-02-23 | 2023-02-23 | 2023-02-24 |
| B | 2023-02-27 | 2023-02-27 | 2023-03-01 |
| B | 2023-02-28 | 2023-02-28 | 2023-03-01 |
| B | 2023-02-28 | 2023-02-28 | 2023-03-01 |
| B | 2023-03-01 | 2023-03-01 | 2023-03-03 |
| B | 2023-03-01 | 2023-03-01 | 2023-03-06 |
| B | 2023-03-02 | 2023-03-02 | 2023-03-06 |
| B | 2023-03-03 | 2023-03-03 | 2023-03-08 |
| B | 2023-03-06 | 2023-03-06 | 2023-03-07 |
| B | 2023-03-08 | 2023-03-08 | 2023-03-09 |

### D Molecular line list data

Table S3 shows the line list data for the laboratory test results from human saliva samples. For each class and test, it shows the date when the test was taken, the test result and (if positive) the virus that was detected. Recall that all students in class were sampled twice per week every Tuesday and Thursday.

**Table S3.** Line list of respiratory cases over the study period. IFB: Influenza B, HRV: rhinovirus, AdV: adenovirus, CoV: SARS-CoV-2, MPV: metapneumovirus, PIV: parainfluenza virus.

| Class | Date of test | Weekday | Test result | Virus |
|---|---|---|---|---|
| A | 2023-01-17 | Tuesday | Negative | - |
| A | 2023-01-17 | Tuesday | Negative | - |
| A | 2023-01-17 | Tuesday | Negative | - |
| A | 2023-01-17 | Tuesday | Negative | - |
| A | 2023-01-17 | Tuesday | Negative | - |
| A | 2023-01-17 | Tuesday | Negative | - |
| A | 2023-01-17 | Tuesday | Negative | - |
| A | 2023-01-17 | Tuesday | Negative | - |
| A | 2023-01-17 | Tuesday | Negative | - |
| A | 2023-01-17 | Tuesday | Negative | - |
| A | 2023-01-17 | Tuesday | Negative | - |
| A | 2023-01-17 | Tuesday | Negative | - |
| A | 2023-01-17 | Tuesday | Negative | - |
| A | 2023-01-17 | Tuesday | Negative | - |
| A | 2023-01-17 | Tuesday | Negative | - |
| A | 2023-01-17 | Tuesday | Positive | AdV |
| A | 2023-01-17 | Tuesday | Positive | AdV |
| A | 2023-01-19 | Thursday | Negative | - |
| A | 2023-01-19 | Thursday | Negative | - |
| A | 2023-01-19 | Thursday | Negative | - |
| A | 2023-01-19 | Thursday | Negative | - |
| A | 2023-01-19 | Thursday | Negative | - |
| A | 2023-01-19 | Thursday | Negative | - |
| A | 2023-01-19 | Thursday | Negative | - |
| A | 2023-01-19 | Thursday | Negative | - |
| A | 2023-01-19 | Thursday | Negative | - |

| | | | | |
|---|---|---|---|---|
| A | 2023-01-19 | Thursday | Negative | - |
| A | 2023-01-19 | Thursday | Negative | - |
| A | 2023-01-19 | Thursday | Negative | - |
| A | 2023-01-19 | Thursday | Negative | - |
| A | 2023-01-19 | Thursday | Negative | - |
| A | 2023-01-19 | Thursday | Positive | AdV |
| A | 2023-01-19 | Thursday | Positive | AdV |
| A | 2023-01-24 | Tuesday | Negative | - |
| A | 2023-01-24 | Tuesday | Negative | - |
| A | 2023-01-24 | Tuesday | Negative | - |
| A | 2023-01-24 | Tuesday | Negative | - |
| A | 2023-01-24 | Tuesday | Negative | - |
| A | 2023-01-24 | Tuesday | Negative | - |
| A | 2023-01-24 | Tuesday | Negative | - |
| A | 2023-01-24 | Tuesday | Negative | - |
| A | 2023-01-24 | Tuesday | Negative | - |
| A | 2023-01-24 | Tuesday | Negative | - |
| A | 2023-01-24 | Tuesday | Negative | - |
| A | 2023-01-24 | Tuesday | Negative | - |
| A | 2023-01-24 | Tuesday | Negative | - |
| A | 2023-01-24 | Tuesday | Negative | - |
| A | 2023-01-24 | Tuesday | Positive | AdV |
| A | 2023-01-24 | Tuesday | Positive | AdV |
| A | 2023-01-24 | Tuesday | Positive | HRV |
| A | 2023-01-26 | Thursday | Negative | - |
| A | 2023-01-26 | Thursday | Negative | - |
| A | 2023-01-26 | Thursday | Negative | - |
| A | 2023-01-26 | Thursday | Negative | - |
| A | 2023-01-26 | Thursday | Negative | - |
| A | 2023-01-26 | Thursday | Negative | - |
| A | 2023-01-26 | Thursday | Negative | - |
| A | 2023-01-26 | Thursday | Negative | - |
| A | 2023-01-26 | Thursday | Negative | - |
| A | 2023-01-26 | Thursday | Negative | - |
| A | 2023-01-26 | Thursday | Negative | - |
| A | 2023-01-26 | Thursday | Negative | - |

| | | | | |
|---|---|---|---|---|
| A | 2023-01-26 | Thursday | Negative | - |
| A | 2023-01-26 | Thursday | Negative | - |
| A | 2023-01-26 | Thursday | Negative | - |
| A | 2023-01-26 | Thursday | Positive | AdV |
| A | 2023-01-26 | Thursday | Positive | AdV |
| A | 2023-01-31 | Tuesday | Negative | - |
| A | 2023-01-31 | Tuesday | Negative | - |
| A | 2023-01-31 | Tuesday | Negative | - |
| A | 2023-01-31 | Tuesday | Negative | - |
| A | 2023-01-31 | Tuesday | Negative | - |
| A | 2023-01-31 | Tuesday | Negative | - |
| A | 2023-01-31 | Tuesday | Negative | - |
| A | 2023-01-31 | Tuesday | Negative | - |
| A | 2023-01-31 | Tuesday | Negative | - |
| A | 2023-01-31 | Tuesday | Negative | - |
| A | 2023-01-31 | Tuesday | Negative | - |
| A | 2023-01-31 | Tuesday | Negative | - |
| A | 2023-01-31 | Tuesday | Negative | - |
| A | 2023-01-31 | Tuesday | Negative | - |
| A | 2023-01-31 | Tuesday | Negative | - |
| A | 2023-01-31 | Tuesday | Negative | - |
| A | 2023-01-31 | Tuesday | Negative | - |
| A | 2023-01-31 | Tuesday | Positive | AdV |
| A | 2023-02-02 | Thursday | Negative | - |
| A | 2023-02-02 | Thursday | Negative | - |
| A | 2023-02-02 | Thursday | Negative | - |
| A | 2023-02-02 | Thursday | Negative | - |
| A | 2023-02-02 | Thursday | Negative | - |
| A | 2023-02-02 | Thursday | Negative | - |
| A | 2023-02-02 | Thursday | Negative | - |
| A | 2023-02-02 | Thursday | Negative | - |
| A | 2023-02-02 | Thursday | Negative | - |
| A | 2023-02-02 | Thursday | Negative | - |
| A | 2023-02-02 | Thursday | Negative | - |
| A | 2023-02-02 | Thursday | Negative | - |
| A | 2023-02-02 | Thursday | Negative | - |

| | | | | |
|---|---|---|---|---|
| A | 2023-02-02 | Thursday | Negative | - |
| A | 2023-02-02 | Thursday | Positive | AdV |
| A | 2023-02-02 | Thursday | Positive | AdV |
| A | 2023-02-14 | Tuesday | Negative | - |
| A | 2023-02-14 | Tuesday | Negative | - |
| A | 2023-02-14 | Tuesday | Negative | - |
| A | 2023-02-14 | Tuesday | Negative | - |
| A | 2023-02-14 | Tuesday | Negative | - |
| A | 2023-02-14 | Tuesday | Negative | - |
| A | 2023-02-14 | Tuesday | Negative | - |
| A | 2023-02-14 | Tuesday | Negative | - |
| A | 2023-02-14 | Tuesday | Negative | - |
| A | 2023-02-14 | Tuesday | Negative | - |
| A | 2023-02-14 | Tuesday | Negative | - |
| A | 2023-02-14 | Tuesday | Negative | - |
| A | 2023-02-14 | Tuesday | Negative | - |
| A | 2023-02-14 | Tuesday | Negative | - |
| A | 2023-02-14 | Tuesday | Negative | - |
| A | 2023-02-14 | Tuesday | Negative | - |
| A | 2023-02-14 | Tuesday | Negative | - |
| A | 2023-02-14 | Tuesday | Positive | MPV |
| A | 2023-02-16 | Thursday | Negative | - |
| A | 2023-02-16 | Thursday | Negative | - |
| A | 2023-02-16 | Thursday | Negative | - |
| A | 2023-02-16 | Thursday | Negative | - |
| A | 2023-02-16 | Thursday | Negative | - |
| A | 2023-02-16 | Thursday | Negative | - |
| A | 2023-02-16 | Thursday | Negative | - |
| A | 2023-02-16 | Thursday | Negative | - |
| A | 2023-02-16 | Thursday | Negative | - |
| A | 2023-02-16 | Thursday | Negative | - |
| A | 2023-02-16 | Thursday | Negative | - |
| A | 2023-02-16 | Thursday | Negative | - |
| A | 2023-02-16 | Thursday | Negative | - |
| A | 2023-02-16 | Thursday | Negative | - |
| A | 2023-02-16 | Thursday | Negative | - |

| | | | | |
|---|---|---|---|---|
| A | 2023-02-16 | Thursday | Negative | - |
| A | 2023-02-16 | Thursday | Negative | - |
| A | 2023-02-16 | Thursday | Positive | MPV |
| A | 2023-02-21 | Tuesday | Negative | - |
| A | 2023-02-21 | Tuesday | Negative | - |
| A | 2023-02-21 | Tuesday | Negative | - |
| A | 2023-02-21 | Tuesday | Negative | - |
| A | 2023-02-21 | Tuesday | Negative | - |
| A | 2023-02-21 | Tuesday | Negative | - |
| A | 2023-02-21 | Tuesday | Negative | - |
| A | 2023-02-21 | Tuesday | Negative | - |
| A | 2023-02-21 | Tuesday | Negative | - |
| A | 2023-02-21 | Tuesday | Negative | - |
| A | 2023-02-21 | Tuesday | Negative | - |
| A | 2023-02-21 | Tuesday | Negative | - |
| A | 2023-02-21 | Tuesday | Negative | - |
| A | 2023-02-21 | Tuesday | Negative | - |
| A | 2023-02-21 | Tuesday | Negative | - |
| A | 2023-02-21 | Tuesday | Positive | CoV |
| A | 2023-02-23 | Thursday | Negative | - |
| A | 2023-02-23 | Thursday | Negative | - |
| A | 2023-02-23 | Thursday | Negative | - |
| A | 2023-02-23 | Thursday | Negative | - |
| A | 2023-02-23 | Thursday | Negative | - |
| A | 2023-02-23 | Thursday | Negative | - |
| A | 2023-02-23 | Thursday | Negative | - |
| A | 2023-02-23 | Thursday | Negative | - |
| A | 2023-02-23 | Thursday | Negative | - |
| A | 2023-02-23 | Thursday | Negative | - |
| A | 2023-02-23 | Thursday | Negative | - |
| A | 2023-02-23 | Thursday | Negative | - |
| A | 2023-02-23 | Thursday | Negative | - |
| A | 2023-02-23 | Thursday | Negative | - |
| A | 2023-02-23 | Thursday | Negative | - |
| A | 2023-02-23 | Thursday | Negative | - |
| A | 2023-02-28 | Tuesday | Negative | - |

| | | | | |
|---|---|---|---|---|
| A | 2023-02-28 | Tuesday | Negative | - |
| A | 2023-02-28 | Tuesday | Negative | - |
| A | 2023-02-28 | Tuesday | Negative | - |
| A | 2023-02-28 | Tuesday | Negative | - |
| A | 2023-02-28 | Tuesday | Negative | - |
| A | 2023-02-28 | Tuesday | Negative | - |
| A | 2023-02-28 | Tuesday | Negative | - |
| A | 2023-02-28 | Tuesday | Negative | - |
| A | 2023-02-28 | Tuesday | Negative | - |
| A | 2023-02-28 | Tuesday | Negative | - |
| A | 2023-02-28 | Tuesday | Negative | - |
| A | 2023-02-28 | Tuesday | Negative | - |
| A | 2023-02-28 | Tuesday | Positive | IFB |
| A | 2023-03-02 | Thursday | Negative | - |
| A | 2023-03-02 | Thursday | Negative | - |
| A | 2023-03-02 | Thursday | Negative | - |
| A | 2023-03-02 | Thursday | Negative | - |
| A | 2023-03-02 | Thursday | Negative | - |
| A | 2023-03-02 | Thursday | Negative | - |
| A | 2023-03-02 | Thursday | Negative | - |
| A | 2023-03-02 | Thursday | Negative | - |
| A | 2023-03-02 | Thursday | Negative | - |
| A | 2023-03-02 | Thursday | Negative | - |
| A | 2023-03-02 | Thursday | Negative | - |
| A | 2023-03-02 | Thursday | Negative | - |
| A | 2023-03-02 | Thursday | Negative | - |
| A | 2023-03-02 | Thursday | Negative | - |
| A | 2023-03-02 | Thursday | Positive | AdV |
| A | 2023-03-07 | Tuesday | Negative | - |
| A | 2023-03-07 | Tuesday | Negative | - |
| A | 2023-03-07 | Tuesday | Negative | - |
| A | 2023-03-07 | Tuesday | Negative | - |
| A | 2023-03-07 | Tuesday | Negative | - |
| A | 2023-03-07 | Tuesday | Negative | - |
| A | 2023-03-07 | Tuesday | Negative | - |
| A | 2023-03-07 | Tuesday | Negative | - |

| | | | | |
|---|---|---|---|---|
| A | 2023-03-07 | Tuesday | Negative | - |
| A | 2023-03-07 | Tuesday | Negative | - |
| A | 2023-03-07 | Tuesday | Negative | - |
| A | 2023-03-07 | Tuesday | Negative | - |
| A | 2023-03-07 | Tuesday | Negative | - |
| A | 2023-03-07 | Tuesday | Negative | - |
| A | 2023-03-07 | Tuesday | Negative | - |
| A | 2023-03-07 | Tuesday | Positive | HRV |
| A | 2023-03-07 | Tuesday | Positive | HRV |
| A | 2023-03-09 | Thursday | Negative | - |
| A | 2023-03-09 | Thursday | Negative | - |
| A | 2023-03-09 | Thursday | Negative | - |
| A | 2023-03-09 | Thursday | Negative | - |
| A | 2023-03-09 | Thursday | Negative | - |
| A | 2023-03-09 | Thursday | Negative | - |
| A | 2023-03-09 | Thursday | Negative | - |
| A | 2023-03-09 | Thursday | Negative | - |
| A | 2023-03-09 | Thursday | Negative | - |
| A | 2023-03-09 | Thursday | Negative | - |
| A | 2023-03-09 | Thursday | Negative | - |
| A | 2023-03-09 | Thursday | Negative | - |
| A | 2023-03-09 | Thursday | Negative | - |
| A | 2023-03-09 | Thursday | Positive | HRV |
| A | 2023-03-09 | Thursday | Positive | HRV |
| A | 2023-03-09 | Thursday | Positive | HRV |
| A | 2023-03-09 | Thursday | Positive | IFB |
| B | 2023-01-17 | Tuesday | Negative | - |
| B | 2023-01-17 | Tuesday | Negative | - |
| B | 2023-01-17 | Tuesday | Negative | - |
| B | 2023-01-17 | Tuesday | Negative | - |
| B | 2023-01-17 | Tuesday | Negative | - |
| B | 2023-01-17 | Tuesday | Negative | - |
| B | 2023-01-17 | Tuesday | Negative | - |
| B | 2023-01-17 | Tuesday | Negative | - |
| B | 2023-01-17 | Tuesday | Negative | - |
| B | 2023-01-17 | Tuesday | Negative | - |

| | | | | |
|---|---|---|---|---|
| B | 2023-01-17 | Tuesday | Negative | - |
| B | 2023-01-17 | Tuesday | Negative | - |
| B | 2023-01-17 | Tuesday | Positive | HRV |
| B | 2023-01-17 | Tuesday | Positive | HRV |
| B | 2023-01-17 | Tuesday | Positive | IFB |
| B | 2023-01-19 | Thursday | Negative | - |
| B | 2023-01-19 | Thursday | Negative | - |
| B | 2023-01-19 | Thursday | Negative | - |
| B | 2023-01-19 | Thursday | Negative | - |
| B | 2023-01-19 | Thursday | Negative | - |
| B | 2023-01-19 | Thursday | Negative | - |
| B | 2023-01-19 | Thursday | Negative | - |
| B | 2023-01-19 | Thursday | Negative | - |
| B | 2023-01-19 | Thursday | Negative | - |
| B | 2023-01-19 | Thursday | Negative | - |
| B | 2023-01-19 | Thursday | Negative | - |
| B | 2023-01-19 | Thursday | Negative | - |
| B | 2023-01-19 | Thursday | Negative | - |
| B | 2023-01-19 | Thursday | Negative | - |
| B | 2023-01-19 | Thursday | Negative | - |
| B | 2023-01-19 | Thursday | Positive | IFB |
| B | 2023-01-24 | Tuesday | Negative | - |
| B | 2023-01-24 | Tuesday | Negative | - |
| B | 2023-01-24 | Tuesday | Negative | - |
| B | 2023-01-24 | Tuesday | Negative | - |
| B | 2023-01-24 | Tuesday | Negative | - |
| B | 2023-01-24 | Tuesday | Negative | - |
| B | 2023-01-24 | Tuesday | Negative | - |
| B | 2023-01-24 | Tuesday | Negative | - |
| B | 2023-01-24 | Tuesday | Negative | - |
| B | 2023-01-24 | Tuesday | Negative | - |
| B | 2023-01-24 | Tuesday | Negative | - |
| B | 2023-01-24 | Tuesday | Negative | - |
| B | 2023-01-24 | Tuesday | Positive | HRV |
| B | 2023-01-26 | Thursday | Negative | - |
| B | 2023-01-26 | Thursday | Negative | - |

| | | | | |
|---|---|---|---|---|
| B | 2023-01-26 | Thursday | Negative | - |
| B | 2023-01-26 | Thursday | Negative | - |
| B | 2023-01-26 | Thursday | Negative | - |
| B | 2023-01-26 | Thursday | Negative | - |
| B | 2023-01-26 | Thursday | Negative | - |
| B | 2023-01-26 | Thursday | Negative | - |
| B | 2023-01-26 | Thursday | Negative | - |
| B | 2023-01-26 | Thursday | Negative | - |
| B | 2023-01-26 | Thursday | Negative | - |
| B | 2023-01-26 | Thursday | Negative | - |
| B | 2023-01-26 | Thursday | Positive | IFB |
| B | 2023-01-26 | Thursday | Positive | IFB |
| B | 2023-01-26 | Thursday | Positive | PIV |
| B | 2023-01-31 | Tuesday | Negative | - |
| B | 2023-01-31 | Tuesday | Negative | - |
| B | 2023-01-31 | Tuesday | Negative | - |
| B | 2023-01-31 | Tuesday | Negative | - |
| B | 2023-01-31 | Tuesday | Negative | - |
| B | 2023-01-31 | Tuesday | Negative | - |
| B | 2023-01-31 | Tuesday | Negative | - |
| B | 2023-01-31 | Tuesday | Negative | - |
| B | 2023-01-31 | Tuesday | Negative | - |
| B | 2023-01-31 | Tuesday | Negative | - |
| B | 2023-01-31 | Tuesday | Negative | - |
| B | 2023-01-31 | Tuesday | Negative | - |
| B | 2023-01-31 | Tuesday | Negative | - |
| B | 2023-01-31 | Tuesday | Negative | - |
| B | 2023-01-31 | Tuesday | Positive | IFB |
| B | 2023-01-31 | Tuesday | Positive | IFB |
| B | 2023-02-02 | Thursday | Negative | - |
| B | 2023-02-02 | Thursday | Negative | - |
| B | 2023-02-02 | Thursday | Negative | - |
| B | 2023-02-02 | Thursday | Negative | - |
| B | 2023-02-02 | Thursday | Negative | - |
| B | 2023-02-02 | Thursday | Negative | - |
| B | 2023-02-02 | Thursday | Negative | - |

| | | | | |
|---|---|---|---|---|
| B | 2023-02-02 | Thursday | Negative | - |
| B | 2023-02-02 | Thursday | Negative | - |
| B | 2023-02-02 | Thursday | Negative | - |
| B | 2023-02-02 | Thursday | Negative | - |
| B | 2023-02-02 | Thursday | Negative | - |
| B | 2023-02-02 | Thursday | Negative | - |
| B | 2023-02-02 | Thursday | Negative | - |
| B | 2023-02-02 | Thursday | Negative | - |
| B | 2023-02-02 | Thursday | Positive | IFB |
| B | 2023-02-14 | Tuesday | Negative | - |
| B | 2023-02-14 | Tuesday | Negative | - |
| B | 2023-02-14 | Tuesday | Negative | - |
| B | 2023-02-14 | Tuesday | Negative | - |
| B | 2023-02-14 | Tuesday | Negative | - |
| B | 2023-02-14 | Tuesday | Negative | - |
| B | 2023-02-14 | Tuesday | Negative | - |
| B | 2023-02-14 | Tuesday | Negative | - |
| B | 2023-02-14 | Tuesday | Negative | - |
| B | 2023-02-14 | Tuesday | Negative | - |
| B | 2023-02-14 | Tuesday | Negative | - |
| B | 2023-02-14 | Tuesday | Negative | - |
| B | 2023-02-14 | Tuesday | Negative | - |
| B | 2023-02-14 | Tuesday | Negative | - |
| B | 2023-02-14 | Tuesday | Negative | - |
| B | 2023-02-14 | Tuesday | Negative | - |
| B | 2023-02-14 | Tuesday | Negative | - |
| B | 2023-02-14 | Tuesday | Positive | IFB |
| B | 2023-02-16 | Thursday | Negative | - |
| B | 2023-02-16 | Thursday | Negative | - |
| B | 2023-02-16 | Thursday | Negative | - |
| B | 2023-02-16 | Thursday | Negative | - |
| B | 2023-02-16 | Thursday | Negative | - |
| B | 2023-02-16 | Thursday | Negative | - |
| B | 2023-02-16 | Thursday | Negative | - |
| B | 2023-02-16 | Thursday | Negative | - |
| B | 2023-02-16 | Thursday | Negative | - |

| | | | | |
|---|---|---|---|---|
| B | 2023-02-16 | Thursday | Negative | - |
| B | 2023-02-16 | Thursday | Negative | - |
| B | 2023-02-16 | Thursday | Negative | - |
| B | 2023-02-16 | Thursday | Negative | - |
| B | 2023-02-16 | Thursday | Negative | - |
| B | 2023-02-16 | Thursday | Negative | - |
| B | 2023-02-16 | Thursday | Negative | - |
| B | 2023-02-16 | Thursday | Negative | - |
| B | 2023-02-16 | Thursday | Negative | - |
| B | 2023-02-21 | Tuesday | Negative | - |
| B | 2023-02-21 | Tuesday | Negative | - |
| B | 2023-02-21 | Tuesday | Negative | - |
| B | 2023-02-21 | Tuesday | Negative | - |
| B | 2023-02-21 | Tuesday | Negative | - |
| B | 2023-02-21 | Tuesday | Negative | - |
| B | 2023-02-21 | Tuesday | Negative | - |
| B | 2023-02-21 | Tuesday | Negative | - |
| B | 2023-02-21 | Tuesday | Negative | - |
| B | 2023-02-21 | Tuesday | Negative | - |
| B | 2023-02-21 | Tuesday | Negative | - |
| B | 2023-02-21 | Tuesday | Negative | - |
| B | 2023-02-21 | Tuesday | Negative | - |
| B | 2023-02-21 | Tuesday | Negative | - |
| B | 2023-02-21 | Tuesday | Positive | CoV |
| B | 2023-02-21 | Tuesday | Positive | HRV |
| B | 2023-02-23 | Thursday | Negative | - |
| B | 2023-02-23 | Thursday | Negative | - |
| B | 2023-02-23 | Thursday | Negative | - |
| B | 2023-02-23 | Thursday | Negative | - |
| B | 2023-02-23 | Thursday | Negative | - |
| B | 2023-02-23 | Thursday | Negative | - |
| B | 2023-02-23 | Thursday | Negative | - |
| B | 2023-02-23 | Thursday | Negative | - |
| B | 2023-02-23 | Thursday | Negative | - |
| B | 2023-02-23 | Thursday | Negative | - |
| B | 2023-02-23 | Thursday | Negative | - |

| | | | | |
|---|---|---|---|---|
| B | 2023-02-23 | Thursday | Negative | - |
| B | 2023-02-23 | Thursday | Positive | CoV |
| B | 2023-02-23 | Thursday | Positive | HRV |
| B | 2023-02-28 | Tuesday | Negative | - |
| B | 2023-02-28 | Tuesday | Negative | - |
| B | 2023-02-28 | Tuesday | Negative | - |
| B | 2023-02-28 | Tuesday | Negative | - |
| B | 2023-02-28 | Tuesday | Negative | - |
| B | 2023-02-28 | Tuesday | Negative | - |
| B | 2023-02-28 | Tuesday | Negative | - |
| B | 2023-02-28 | Tuesday | Negative | - |
| B | 2023-02-28 | Tuesday | Negative | - |
| B | 2023-02-28 | Tuesday | Negative | - |
| B | 2023-02-28 | Tuesday | Negative | - |
| B | 2023-02-28 | Tuesday | Positive | HRV |
| B | 2023-02-28 | Tuesday | Positive | IFB |
| B | 2023-03-02 | Thursday | Negative | - |
| B | 2023-03-02 | Thursday | Negative | - |
| B | 2023-03-02 | Thursday | Negative | - |
| B | 2023-03-02 | Thursday | Negative | - |
| B | 2023-03-02 | Thursday | Negative | - |
| B | 2023-03-02 | Thursday | Negative | - |
| B | 2023-03-02 | Thursday | Negative | - |
| B | 2023-03-02 | Thursday | Negative | - |
| B | 2023-03-02 | Thursday | Negative | - |
| B | 2023-03-02 | Thursday | Negative | - |
| B | 2023-03-02 | Thursday | Negative | - |
| B | 2023-03-02 | Thursday | Positive | IFB |
| B | 2023-03-07 | Tuesday | Negative | - |
| B | 2023-03-07 | Tuesday | Negative | - |
| B | 2023-03-07 | Tuesday | Negative | - |
| B | 2023-03-07 | Tuesday | Negative | - |
| B | 2023-03-07 | Tuesday | Negative | - |
| B | 2023-03-07 | Tuesday | Negative | - |
| B | 2023-03-07 | Tuesday | Negative | - |
| B | 2023-03-07 | Tuesday | Negative | - |

| | | | | |
|---|---|---|---|---|
| B | 2023-03-07 | Tuesday | Negative | - |
| B | 2023-03-07 | Tuesday | Negative | - |
| B | 2023-03-07 | Tuesday | Negative | - |
| B | 2023-03-07 | Tuesday | Negative | - |
| B | 2023-03-07 | Tuesday | Negative | - |
| B | 2023-03-07 | Tuesday | Positive | AdV |
| B | 2023-03-07 | Tuesday | Positive | HRV |
| B | 2023-03-07 | Tuesday | Positive | IFB |
| B | 2023-03-07 | Tuesday | Positive | IFB |
| B | 2023-03-09 | Thursday | Negative | - |
| B | 2023-03-09 | Thursday | Negative | - |
| B | 2023-03-09 | Thursday | Negative | - |
| B | 2023-03-09 | Thursday | Negative | - |
| B | 2023-03-09 | Thursday | Negative | - |
| B | 2023-03-09 | Thursday | Negative | - |
| B | 2023-03-09 | Thursday | Negative | - |
| B | 2023-03-09 | Thursday | Negative | - |
| B | 2023-03-09 | Thursday | Negative | - |
| B | 2023-03-09 | Thursday | Negative | - |
| B | 2023-03-09 | Thursday | Negative | - |
| B | 2023-03-09 | Thursday | Negative | - |
| B | 2023-03-09 | Thursday | Negative | - |
| B | 2023-03-09 | Thursday | Positive | AdV |
| B | 2023-03-09 | Thursday | Positive | HRV |
| B | 2023-03-09 | Thursday | Positive | HRV |
| B | 2023-03-09 | Thursday | Positive | IFB |

### E Modeling changes in particle concentrations

We only analyze particle concentration data of the time periods during which students were in the classroom. Particle concentrations $Y$ on day $t$ in class $j$ are summarized daily with the mean across these time periods. The change in aerosol number and particle matter mass concentrations is then estimated using Bayesian log-linear regression models

$$\begin{aligned}
\log Y | \beta_0, \beta_1, \ldots, \beta_6 &\sim \text{Normal}(\mu, \sigma) \\
\mu &= \beta_0 + \beta_1 \cdot \text{AirCleaner} + \beta_2 \cdot \text{Class} + \beta_3 \cdot \text{Weekday} + \beta_4 \cdot \text{Students} \\
&\quad + \beta_5 \cdot \text{AirChangeRate} + \beta_6 \cdot \text{Cases} \\
\beta_0 &\sim \text{Student-t}_5(\mu_y, 2.5 s_y) \\
\beta_1, \ldots, \beta_6 &\sim \text{Student-t}_5\left(0, 2.5\frac{s_y}{s_x}\right) \\
\sigma &\sim \text{Exponential}\left(\frac{1}{s_y}\right) .
\end{aligned}$$

The effect of air cleaners is adjusted for class- and weekday-specific effects, the number of students in school, the air change rate, and the cumulative number of cases related to respiratory infections. A log transform is applied to all continuous input variables.

### F Modeling relative risk of infection

#### F.1 Overall approach

The overall aim is to estimate the effects of air cleaners on the daily number of respiratory infections. The latter is unobserved/latent and inferred from the daily number of respiratory cases (absences related to respiratory infections by date of symptom onset), considering the delay from infection to symptom onset (i. e. the incubation period). The effect of air cleaners is estimated with a Bayesian approach, which requires the specification of prior distributions for all model parameters.

#### F.2 Notation

| | |
|---|---|
| $j$ | class |
| $t$ | days since start of study period |
| $C_{jt}$ | number of new cases among students in class $j$ at day $t$ (observed) |
| $I_{jt}$ | number of new infections in class $j$ at day $t$ (unobserved) |
| $F_{jt}$ | number of new infections in class $j$ before day $t$ (unobserved) |
| $N_{jt}$ | cumulative number of infections in class $j$ until day $t$ (unobserved) |

#### F.3 Relating the number of respiratory cases to the number of infections

The number of respiratory cases $C$ in class $j$ at time $t$ is modeled with a Negative Binomial distribution

$$C_{jt} \sim \text{Negative-Binomial}(\mu_{jt}, \phi),$$

where $\mu_{jt}$ is the expected number of new cases and $\phi$ is the parameter modeling over-dispersion. The expected number of new cases is the weighted sum of the number of new infections $I_{jt}$ in the previous days

$$\mu_{jt} = \sum_{s<t} I_{js} \cdot p_{\text{IN}}(t-s),$$

where $p_{\mathrm{IN}}(t-s)$ denotes the probability distribution of the incubation period.

#### F.4 Relating the number of new infections to the presence of interventions

The number of new infections is related to the presence of air cleaners using a log-link

$$\begin{aligned}\log I_{jt} = \log F_{jt} - \log N_{jt} + \beta_0 + \beta_1 \cdot \mathrm{AirCleaner}_{jt} + \beta_2 \cdot \mathrm{Class}_t + \beta_3 \cdot \mathrm{Students}_{jt} \\ + \beta_4 \cdot \mathrm{AirChangeRate}_{jt} + \beta_5 \cdot \mathrm{CoV}_t + \beta_6 \cdot \mathrm{ILI}_t,\end{aligned}$$

where $F_{jt} = \sum_{t-7}^{t-1} I_{js}$ is the number of infections in the previous seven days (first model offset; a proxy for the number of contagious students), $N_{jt} = \sum_{s<t} I_{js}$ is the cumulative number of infections (second model offset; the inverse is a proxy for the number of susceptible students), $\beta_0$ is the rate of new infections without air cleaners (model intercept), and $\beta_1$ is the effect of air cleaners. The effect of air cleaners is adjusted for class-specific effects, the number of students in school, the air change rate, the proportion of positive tests for SARS-CoV-2 in the community, and the number of consultations for influenza-like illnesses in the community.

Note that we deviated from the statistical analysis plan by using as model offset $\log F_{jt} - \log N_{jt}$ instead of just $\log N_{jt}$. This change improved the fit of our model. The reason is that the ratio $\log(F_{jt}/N_{jt})$ incorporates both ways by which new infections can naturally decrease over time, i. e. a decrease in contagious students ($F_{jt}$) or a decrease in susceptible students ($1/N_{jt}$).

#### F.5 Specifying the distribution of the incubation period

The virus of each respiratory infection could not be identified from the epidemiological data because the students never obtained a laboratory test result. As a consequence, different incubation periods need to be considered in $p_{\mathrm{IN}}$, reflecting a combination of the virus-specific incubation periods. The combination is determined based on the weekly proportion of positive saliva samples for each virus found in the molecular analysis. Formally, let $p_{\mathrm{IN}}^{v}$ be the distribution for the incubation period of respiratory virus $v$ and let $pp_{vw}$ be the proportion of positive saliva samples in study week $w$, then each week the combined incubation period is computed as the weighted sum of the virus-specific incubation periods

$$p_{\text{IN}}^{w}(s) = \sum_{v} p_{\text{IN}}^{v}(s) \cdot pp_{vw} \quad \forall s. \tag{3}$$

The prior distributions for the virus-specific incubation periods are based on estimates published in the literature.[6,7] The distributions are shown in Figure S2. Since we could not obtain prior estimates for the incubation period of metapneumovirus (MPV) from the literature, we instead formed a distribution by using the equally weighted average of the parameters from the other distributions.

We estimate the virus-specific distributions from our data as part of fitting the overall model. Furthermore, note that $p_{\text{IN}}$ is discretized via $p_{\text{IN}}(s) = \int_0^{0.5} p_{\text{IN}}(\tau)\,\mathrm{d}\tau$ for $s = 0$ and $p_{\text{IN}}(s) = \int_{s-0.5}^{s+0.5} p_{\text{IN}}(\tau)\,\mathrm{d}\tau$ for $s > 0$, where $p_{\text{IN}}(\tau) \sim \text{Lognormal}(\mu, \sigma)$ is the density of the lognormal distribution.

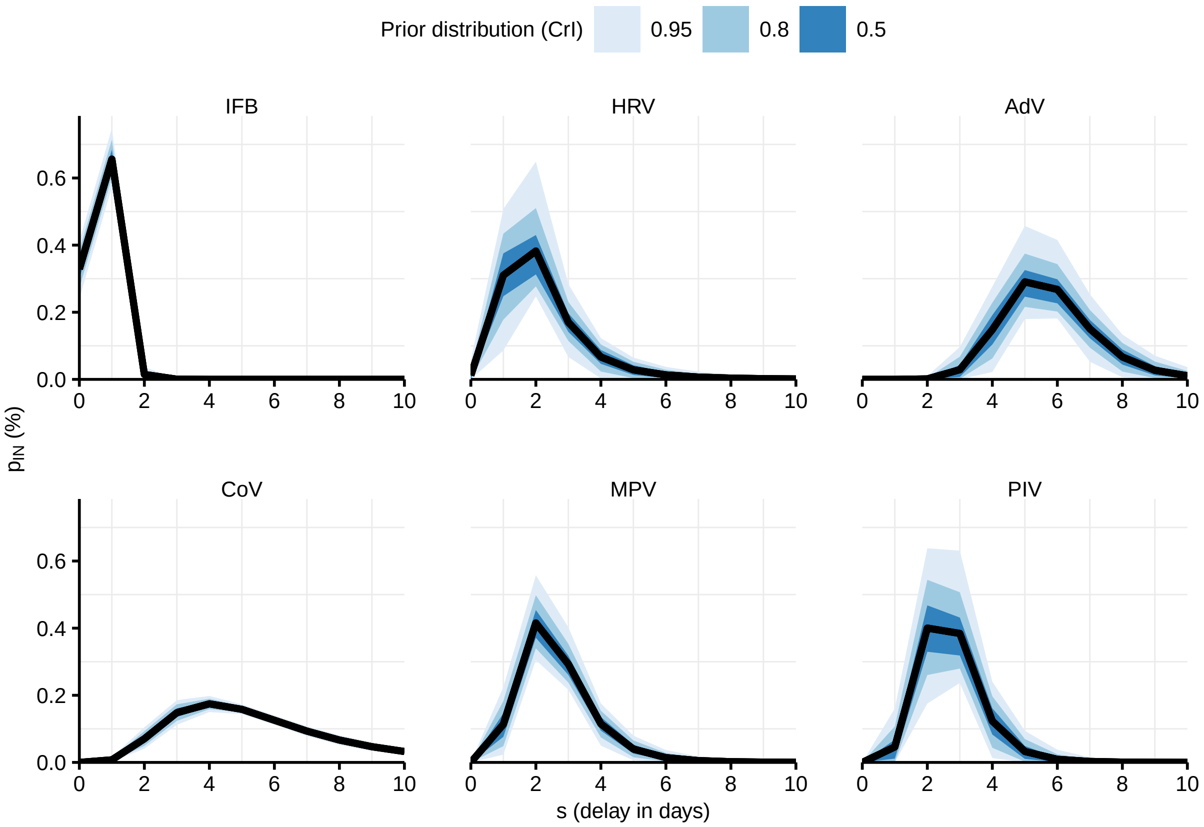

**Figure S2.** Choices of priors for the distribution of the virus-specific incubation periods. IFB: Influenza B, HRV: rhinovirus, AdV: adenovirus, CoV: SARS-CoV-2, MPV: metapneumovirus, PIV: parainfluenza virus.

#### F.6 Adjusting for under-reporting of cases on weekends

Despite recording cases by date of symptom onset, we recorded a higher proportion of cases on Mondays than on weekends, suggesting recall bias and under-reporting of cases on weekends. To consider weekday effects in the reporting of cases, we re-weight the expected number of cases each week as follows. Let $k \in$(1: Saturday, 2: Sunday, 3: Monday, ..., 7: Friday) denote the weekday with the week starting on Saturday. The re-weighted expected number of cases $\tilde{\mu}$ (class and day indexes omitted) are computed as

$$\tilde{\mu}_k = \mu_k \cdot \vartheta_k \cdot \left( \frac{\sum_k \mu_k}{\sum_k \nu_k \cdot \mu_k} \right)$$

$$\sum_k \vartheta_k = 1,$$

where $\nu_k$ is the weight for weekday $k$. These weights are modeled with a Dirichlet prior

$$\vartheta \sim \text{Dirichlet}(c)$$

$$c_k = \sum_j \sum_t C_{jt} \mathbb{I}_{\text{Weekday}(t)=k} \quad \forall k$$

where $c_k$ is the total number of cases reported for weekday $k$ and $\mathbb{I}$ is a binary indicator.

#### F.7 Modeling school-free days

Infections may have occurred during the week of vacation that falls into the study period. The expected number of infections and cases are computed during vacation, but vacation days are not modeled (i. e. not incorporated into the model likelihood). In addition, we assume lower transmission of respiratory infections on days without school (weekends and vacations). We incorporate our prior belief into the model intercept $\beta_0$

$$\beta_0 = \alpha + \omega \cdot \text{NoSchool},$$

where $\alpha$ is the rate of new infections on school days and $\alpha + \omega$ is the rate on days without school. We model $\omega$ with an informative prior for a 10% decrease in new infections on school-free days

$$\omega \sim \text{Normal}(\log 1.1, 0.05) \ .$$

#### F.8 Seeding infections before study start

Cases in the first week of the study could indicate infections before the study commenced. We will therefore seed our model $2 \cdot \mathrm{m}$ days before the study start, where m is the average incubation period of the virus with the largest incubation period (i. e. adenovirus). The number of infections before the study start will be modeled with an exponential prior

$$I_{jt} \sim \text{Exponential}(1) \quad t = -2 \cdot \mathrm{m} + 1, \ldots, 0 \ .$$

Note that we deviated from the statistical analysis plan by changing the parameter of the Exponential distribution from $\lambda = 2 \cdot m$ to $\lambda = 1$. After inspecting the model fit, we realized that the model could not adequately fit the number of new cases at the start of study, because the expected number of new infections in the seeding period $\mu = 1/\lambda = 1/(2 \cdot m)$ were too low. After increasing the expected number of new infections in the seeding period to $\mu = 1$ the model could adequately capture new respiratory cases observed at the study start.

#### F.9 Priors for modeling parameters

We use weakly informative priors for all modeling parameters following recommendations on the choice of priors.[8–12] The continuous adjustment variables are standardized to have zero mean and a standard deviation of 0.5.

$$\begin{aligned} \alpha &\sim \text{Student-t}_5(0, 10) \\ \beta_2, \ldots, \beta_6 &\sim \text{Student-t}_5(0, 2.5) \\ \frac{1}{\sqrt{\phi}} &\sim \text{Half-Normal}(0, 1), \end{aligned}$$

where Student-$\mathrm{t}_5$ is a Student-t distribution with 5 degrees of freedom.

### G Modeling changes in cough frequency

We analyze the daily number of coughs $Y$ with a Negative Binomial regression model

$$\begin{aligned}
Y|\beta_0, \beta_1, \ldots, \beta_6 &\sim \text{Negative-Binomial}(\mu, \phi) \\
\log \mu &= \log T + \beta_0 + \beta_1 \cdot \text{AirCleaner} + \beta_2 \cdot \text{Class} + \beta_3 \cdot \text{Weekday} + \beta_4 \cdot \text{Students} + \beta_5 \cdot \text{Ventilation} + \beta_6 \cdot \\
\beta_0 &\sim \text{Student-t}_5(0, 2.5) \\
\beta_1, \ldots, \beta_6 &\sim \text{Student-t}_5\left(0, \frac{2.5}{s_x}\right) \\
\frac{1}{\sqrt{\phi}} &\sim \text{Half-Normal}(0, 1),
\end{aligned}$$

where $T$ is the daily duration that students were in the classroom and $\phi$ is the parameter modeling over-dispersion in count data.

In addition, we analyze the association between the daily number of coughs $Y$ and the number of positive saliva test resutls for respiratory viruses with a Negative Binomial hierarchical regression model

$$\begin{aligned}
Y|\beta_0, \theta_{[v]}, \theta, \tau &\sim \text{Negative-Binomial}(\mu, \phi) \\
\log \mu &= \log T + \beta_0 + \sum_v \theta_{[v]} \cdot \text{Saliva}_v \\
\beta_0 &\sim \text{Student-t}_5(0, 2.5) \\
\theta_{[\text{IFB}]}, \ldots, \theta_{[\text{PIV}]} &\sim \text{Normal}\,(\theta, \tau) \\
\theta &\sim \text{Student-t}_5\left(0, \frac{2.5}{s_v}\right) \\
\tau &\sim \text{Half-Normal}\,(0, 1) \\
\frac{1}{\sqrt{\phi}} &\sim \text{Half-Normal}(0, 1),
\end{aligned}$$

where $\theta_{[v]}$ is the partially pooled estimate for respiratory virus $v \in$ (IFB, HRV, AdV, CoV, MPV, PIV), $\tau$ estimates variation between viruses, and $s_v$ is the average standard deviation across the count variables of the respiratory viruses.

### H Modeling positivity rate of human saliva samples

Molecular analysis determined which saliva samples were positive for a respiratory virus. Let $p = 0, 1, \ldots, P$ denote the virus, where 0 refers to negative samples and $P$ is the number of different viruses detected over the study. The number of positive bioaerosol and saliva samples $y_p$ in samples of sizes $n$ is analyzed with a Multinomial logistic regression model

$$
\begin{aligned}
y_0, y_1, \cdots, y_P | n, \beta_0, \beta_1, \beta_2, \beta_3 &\sim \text{Multinomial-Logit}(\theta_0, \theta_1, \ldots \theta_P) \\
\theta &= \text{softmax}(\mu) \\
\mu &= \begin{cases} 0, & \text{if } p = 0, \\ \beta_{0p} + \beta_1 \cdot \text{AirCleaner} + \beta_2 \cdot \text{Class} + \beta_3 \cdot \text{Susceptibles}_p, & \text{if } p > 0. \end{cases} \\
\beta_{0p} &\sim \text{Normal}(0, 2.5) \quad \forall p \\
\beta_1, \beta_2, \beta_3 &\sim \text{Student-t}_5\left(0, \frac{2.5}{s_x}\right)
\end{aligned}
$$

where $\text{softmax}(\mu) = \exp(\mu)/\sum\exp(\mu)$ and $s_x$ is the empirical standard deviation of each input variable. The negative test is set as the reference category. Overall variation in positive samples by virus will be modeled with $\beta_{0p}$ and the effect of air cleaners will be modeled with $\beta_1$. The effect of air cleaners is adjusted for class-specific effects and the decreasing number of susceptibles over time (for each virus computed as the number of students minus the total number of positive samples).

Note that we deviated from the statistical analysis plan by estimating $\beta_{0p}$ and $\beta_1$ without a hierarchical prior. First, there was considerable variation in the overall positivity rate of each virus, so that we decided to use unpooled intercepts $\beta_{0p}$ instead of partially pooled intercepts $\beta_{0[p]}$ for each virus. Second, there was insufficient variation in the data to inform partially pooled estimates $\beta_{1[p]}$ for the effects of air cleaners. Therefore, we decided to only estimate the average effect across viruses, i. e. the completely pooled estimate $\beta_1$.

### I Detailed results for changes in particle concentrations

There was a strong difference in particle concentrations between study conditions (Figure S3a). When adjusting for air change rates and multiple other factors, the aerosol number concentration decreased by 76% (95%-CrI 63% to 86%) with air cleaners (Figure S3b and Table S4). The decrease in the concentration of larger particles ($PM_{10}$) was greater than the decrease in the concentration of smaller particles ($PM_1$ to $PM_4$).

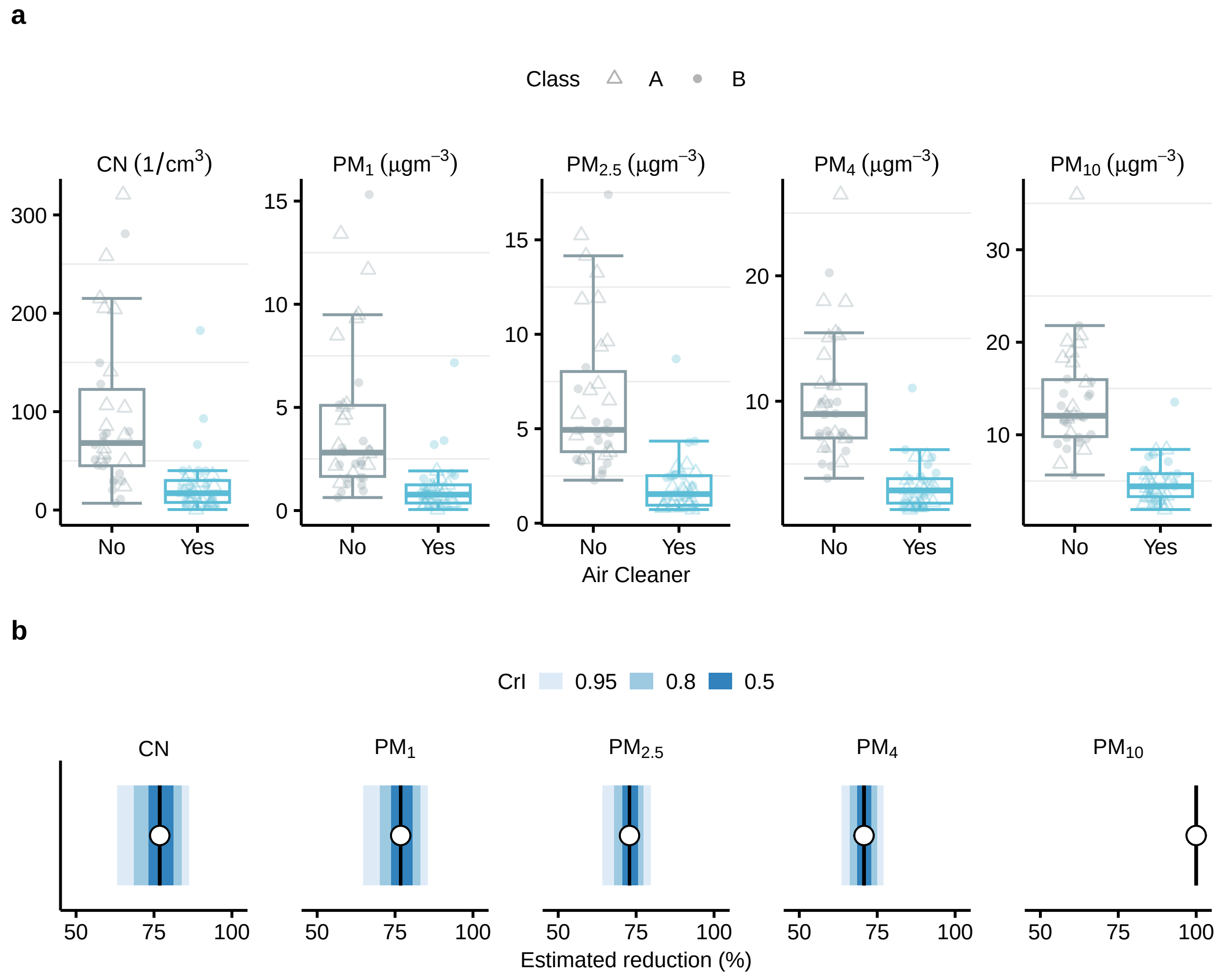

**Figure S3. Analysis of particle concentrations and comparison between study conditions.** **(a)** Boxplot of the daily average values for aerosol number concentration (CN in 1/cm$^3$) and particle mass concentration (PM for particles of sizes <1 to <10 $\mu$m, respectively in $\mu$gm$^{-3}$). **(b)** Estimated reduction in aerosol number and particle mass concentrations with air cleaners (posterior mean as dot and 50%-, 80%- and 95%-CrI as lines, respectively).

In the main paper, we compared particle concentrations between study conditions. Figure S4

**Table S4.** Estimated reduction in aerosol number (CN) and particle mass (PM) concentrations with interventions (posterior mean and upper and lower estimate from the 95%-CrI).

| Variable | Mean | Lower | Upper |
|---|---|---|---|
| CN | 76.83 | 63.20 | 86.28 |
| $PM_1$ | 76.79 | 64.84 | 85.46 |
| $PM_{2.5}$ | 72.86 | 64.24 | 79.66 |
| $PM_4$ | 70.77 | 63.46 | 76.97 |
| $PM_{10}$ | 99.99 | 99.94 | 100.00 |

compares additional environmental variables, i. e. $CO_2$, air change rate, relative humidity, and temperature. The air change rate was estimated from indoor $CO_2$ levels using a transient mass balance model.[13] There are no considerable differences between in any of the variables between study conditions. Furthermore, numerical estimation results for the reduction in particle concentrations with air cleaners are shown in Table S4.

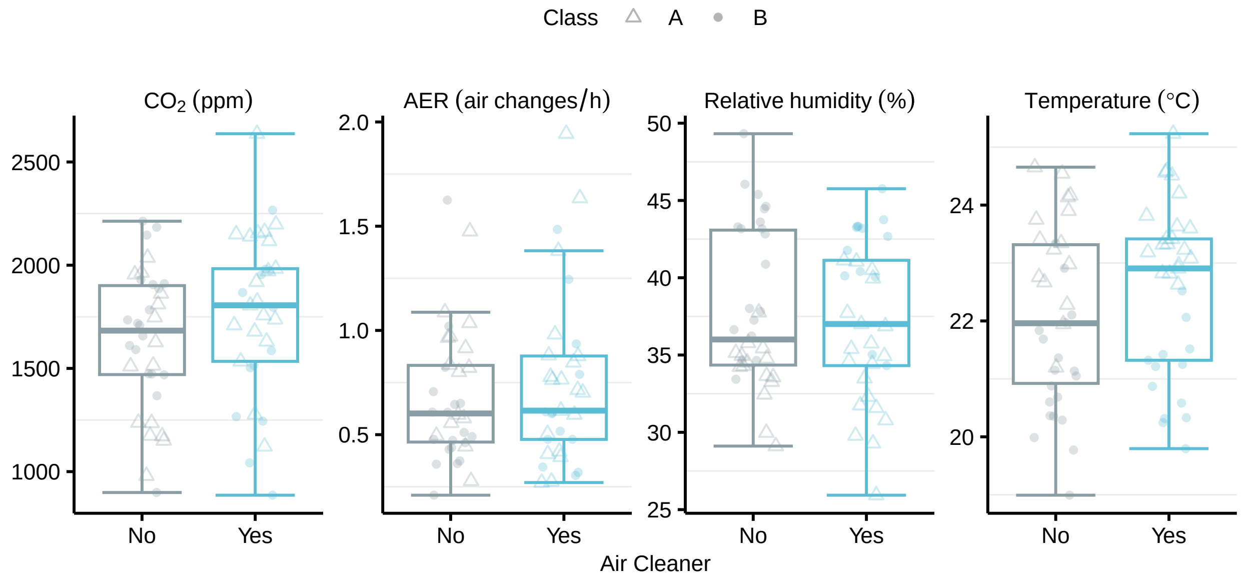

**Figure S4.** Boxplot for the daily average values of each environmental variable by study condition.

### J Detailed results for risk of infection model

Table S5 presents the posterior mean, credible intervals and model diagnostics for all model parameters. See the main paper for a discussion of the effects of interventions. Here we briefly discuss some of the additional model parameters.

The effective sample size (ESS) and the Gelman-Rubin convergence diagnostic ($\hat{R}$) indicate good estimation power. It further suggests that the Markov chains converged.

The estimate for $\beta_2$ is positive, suggesting a higher risk of infection in class $B$, which is in line with the higher number of cases in this class. The estimate for $\beta_3$ is positive, suggesting a higher risk of infection as more students were in class. The estimate for $\beta_4$ is negative, suggesting a lower risk of infection with higher air change rates, in line with the intuition that the risk of infection is lower when the classroom is better ventilated. The estimate for $\beta_5$ and $\beta_6$ are both positive although only the former is distinguishable from zero, suggesting that higher transmission in the community was also associated with a higher risk of infection in the school.

The overdispersion parameter ($\phi$) cannot be precisely estimated, but the credible interval suggests rather small overdispersion ($\phi \to \infty$). The school-free effect $\omega$, the weekday weights $\vartheta$, and the parameters for the distribution of the incubation period $\mu_{p_{\text{IN}}}$ and $\sigma_{p_{\text{IN}}}$ are all close to their priors, suggest that these parameters are barely informed by date but more by our prior assumptions.

Our model accounts for the delay from infection to case confirmation and allows transmission to change only at the dates of interventions. It thus reflects overall trends in transmission during study conditions rather than day-to-day variation. To evaluate how well our model fits these trends, Figure S5 compares the model-estimated expected number of cases with the observed number cases only on a weekly basis and across classes. Overall the estimates are in relatively good agreement. The 95%-CrIs of the model-estimates always include the observed cases.

**Table S5.** Estimation results from infection risk model for the number of new respiratory cases.

| Parameter | Mean | Lower 95%-CrI | Upper 95%-CrI | $\hat{R}$ | ESS |
|---|---|---|---|---|---|
| $\phi$ | 4785.34 | 0.68 | 2733.84 | 1.00 | 2675 |
| $\alpha$ | −0.59 | −1.54 | 0.26 | 1.00 | 2457 |
| $\omega$ | 0.10 | 0.00 | 0.19 | 1.00 | 10312 |
| $\beta_1$ | −0.32 | −0.81 | 0.17 | 1.00 | 2613 |
| $\beta_2$ | 0.92 | −0.04 | 1.92 | 1.00 | 2809 |
| $\beta_3$ | 1.07 | 0.56 | 1.72 | 1.00 | 2183 |
| $\beta_4$ | −1.40 | −2.70 | −0.22 | 1.00 | 1743 |
| $\beta_5$ | 0.81 | 0.48 | 1.15 | 1.00 | 2006 |
| $\beta_6$ | 0.10 | −0.20 | 0.50 | 1.00 | 2089 |
| $\vartheta_{\text{Saturday}}$ | 0.05 | 0.01 | 0.12 | 1.00 | 7401 |
| $\vartheta_{\text{Sunday}}$ | 0.03 | 0.00 | 0.08 | 1.00 | 7481 |
| $\vartheta_{\text{Monday}}$ | 0.47 | 0.34 | 0.60 | 1.00 | 7909 |
| $\vartheta_{\text{Tuesday}}$ | 0.13 | 0.06 | 0.22 | 1.00 | 9220 |
| $\vartheta_{\text{Wednesday}}$ | 0.12 | 0.05 | 0.21 | 1.00 | 6220 |
| $\vartheta_{\text{Thursday}}$ | 0.09 | 0.04 | 0.18 | 1.00 | 6539 |
| $\vartheta_{\text{Friday}}$ | 0.11 | 0.05 | 0.20 | 1.00 | 6028 |
| $\mu_{p_{\text{IN}}^{\text{IFB}}}$ | −0.51 | −0.60 | −0.42 | 1.00 | 8269 |
| $\mu_{p_{\text{IN}}^{\text{HRV}}}$ | 0.76 | 0.50 | 1.02 | 1.00 | 7381 |
| $\mu_{p_{\text{IN}}^{\text{AdV}}}$ | 1.72 | 1.59 | 1.87 | 1.00 | 7370 |
| $\mu_{p_{\text{IN}}^{\text{CoV}}}$ | 1.63 | 1.51 | 1.75 | 1.00 | 9076 |
| $\mu_{p_{\text{IN}}^{\text{MPV}}}$ | 0.89 | 0.73 | 1.05 | 1.00 | 7591 |
| $\mu_{p_{\text{IN}}^{\text{PIV}}}$ | 0.96 | 0.76 | 1.16 | 1.00 | 10017 |
| $\sigma_{p_{\text{IN}}^{\text{IFB}}}$ | 0.42 | 0.33 | 0.50 | 1.00 | 7320 |
| $\sigma_{p_{\text{IN}}^{\text{HRV}}}$ | 0.51 | 0.25 | 0.78 | 1.00 | 3717 |
| $\sigma_{p_{\text{IN}}^{\text{AdV}}}$ | 0.24 | 0.14 | 0.34 | 1.00 | 5458 |
| $\sigma_{p_{\text{IN}}^{\text{CoV}}}$ | 0.50 | 0.45 | 0.54 | 1.00 | 8201 |
| $\sigma_{p_{\text{IN}}^{\text{MPV}}}$ | 0.40 | 0.27 | 0.53 | 1.00 | 7352 |
| $\sigma_{p_{\text{IN}}^{\text{PIV}}}$ | 0.30 | 0.15 | 0.45 | 1.00 | 5224 |

ESS is the effective sample size, i. e. the number of independent MCMC samples with estimation power equivalent to the total number of autocorrelated samples,[14] and $\hat{R}$ is the Gelman-Rubin convergence diagnostic.[15] Low ESS or $\hat{R}$ or $\hat{R} > 1.10$ indicate bad convergence of the model.[16]

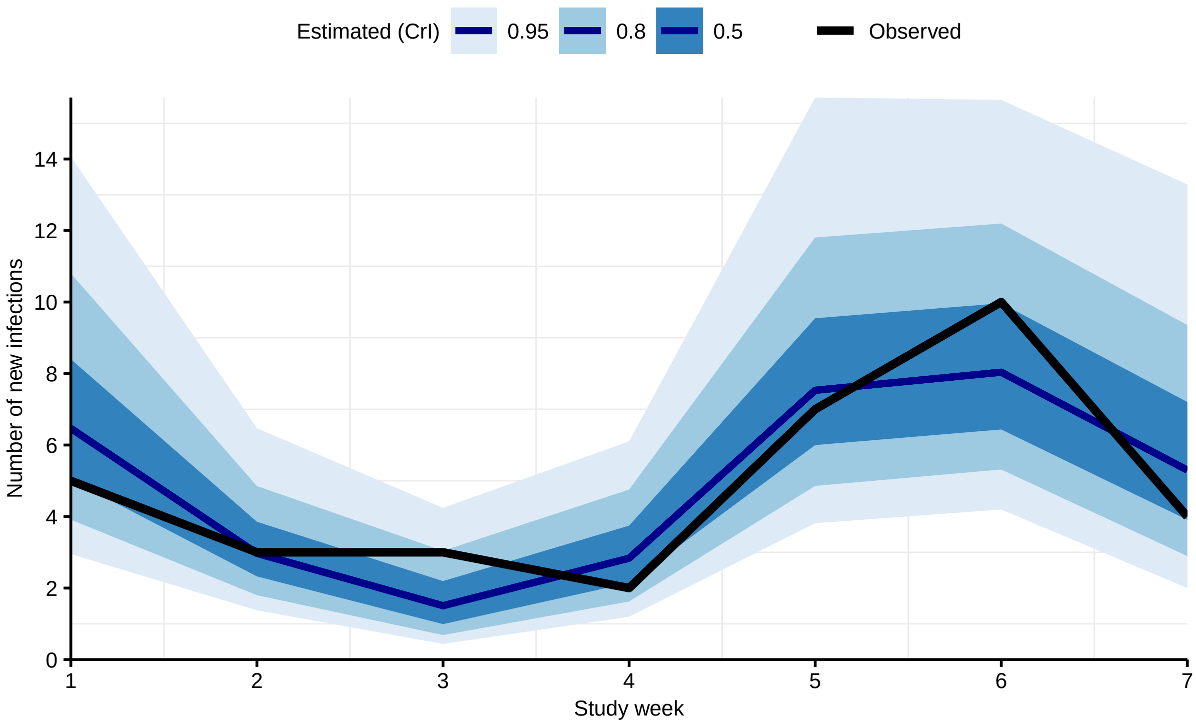

**Figure S5.** Model-estimated number of new respiratory cases (posterior mean as red line and 50%-, 80%, and 95%-CrIs as shaded areas) and the observed number of new respiratory cases (black line) across classes by study week.

### K Detailed results for cough frequency

Differences in the frequency of coughs between classes suggest an association with class-specific virus patterns. Figure S6 shows the relative risk of cough for each respiratory virus. Cough appeared more frequent for IFB and less frequent for AdV.

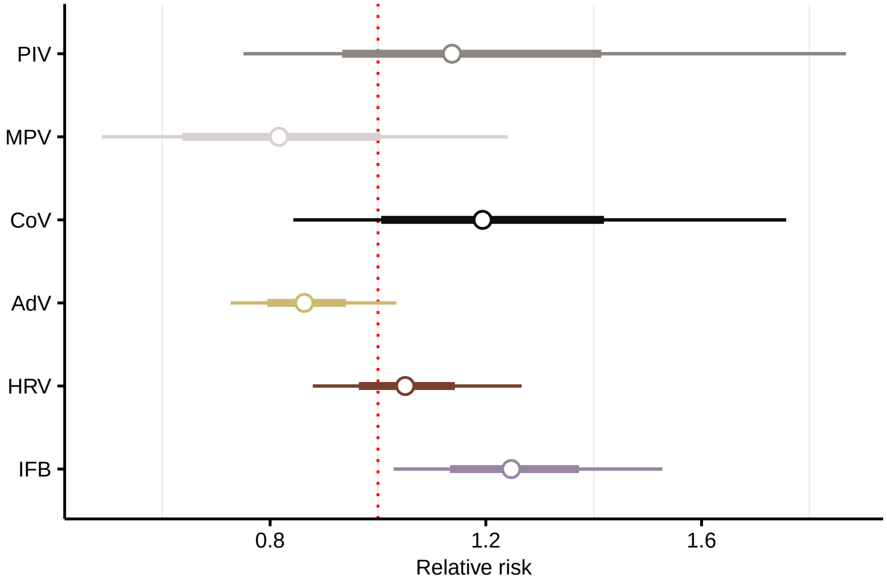

**Figure S6.** Estimated association (relative risk; posterior mean as dot, 50%, 80%, and 95%-CrI as lines) between cough frequency and number of positive saliva test results by respiratory virus. IFB: Influenza B, HRV: rhinovirus, AdV: adenovirus, CoV: SARS-CoV-2, MPV: metapneumovirus, PIV: parainfluenza virus.

### L Detailed results for positivity rate in saliva samples

There can be unknown delay between when students were infected and when they tested positive. Therefore, we performed a sensitivity analysis where we lagged the molecular test results (number of positive saliva samples) by one test date (i. e. from Tuesday to last Thursday and from Thursday to Tuesday) and one test week (i. e. from Tuesday to last Tuesday and Thursday to last Thursday). The results are shown in Figure S7. The estimated effect of air cleaners remains indistinguishable from zero regardless of the considered lag.

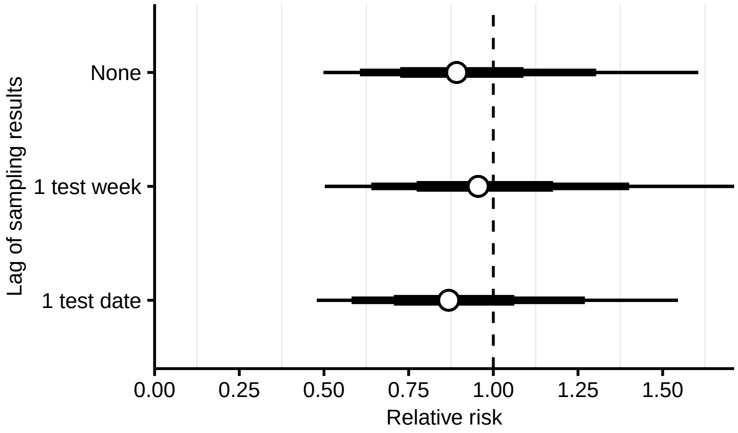

**Figure S7.** Sensitivity analysis for the estimated effect of air cleaners on the positivity rate of saliva samples. Shown is the adjusted relative risk (posterior mean as dot and 50%-, 80%, and 95%-CrIs as lines) for air cleaners by lag of the molecular test results.
